# Predicting PANSS symptoms in schizophrenia spectrum disorders using speech only: an international, multi-centre, computational secondary analysis across multiple languages

**DOI:** 10.64898/2026.02.20.26345632

**Authors:** Rui He, Maryia Kirdun, Claudio Palominos, Lara Navarrete Orejudo, Sophie Barthelemy, Simran Bhola, Silvia Ciampelli, Amandine Decker, Cemal Demirlek, Riccardo Fusaroli, José Tomás García-Molina, Guy Gimenez, Roya Hüppi, Katja Koelkebeck, Amandine Lecomte, Renxiang Qiu, Arndis Simonsen, Vincent Tourneur, Burcu Verim, Huiling Wang, Berna Yalincetin, Sandy Yin, Yuan Zhou, Maxime Amblard, Rosa Ayesa Arriola, Emre Bora, Janna de Boer, Alicia I. Figueroa-Barra, Sanne Koops, Michel Musiol, Lena Palaniyappan, Alberto Parola, Filip Spaniel, Sunny X. Tang, Iris E. Sommer, Philipp Homan, Wolfram Hinzen

## Abstract

**Background:** Speech carries cues to variation in mental state in schizophrenia, typically indexed with clinician-rated scales such as Positive and Negative Syndrome Scale (PANSS). In this study, we aggregate multi-centre recordings to assemble a large corpus and assess symptom-prediction models at scale, to enable rater-independent, efficient, and reproducible assessments for early detection of relapse-related signals from speech.

**Methods:** In this computational secondary analysis, we compiled data from 453 patients with schizophrenia spectrum disorders, recruited between 2005 and 2026 from ten global sites in the Czech Republic, United States, Spain, Chile, France, Switzerland, the Netherlands, Türkiye, Germany, and China, and clipped their speech recordings into 6,664 segments. Across three featuresets, acoustic-prosodic profile, pretrained multilingual embeddings, and their concatenation, we compared 16 algorithms in separate item-specific models to predict eight relapse-related PANSS items, including three positive (P1, P2, P3), three negative (N1, N4, N6), and two general (G5, G9) items, on speaker-disjoint splits. Performance was assessed by root-mean-squared-error (RMSE) at both segment and participant (median-aggregation) levels. Best model per item underwent bias checks for age, sex, education, and symptom severity.

**Findings:** Best-performing models predicted symptoms with prediction errors of 1·5 or lower: P1 1·544/1·527, P2 1·318/1·107, P3 1·487/1·542, N1 1·071/1·030, N4 1·521/1·430, N6 0·869/0·855, G5 0·951/0·882, G9 1·298/1·282 (segment/participant). Performance of pretrained embeddings surpassed acoustic-prosodic features and concatenation. Results were comparable in languages or language variants with fewer available speech and language computational resources. We found no bias by age, sex, or education, aside from reduced N4 accuracy in males; but performance degraded with higher symptom severity.

**Interpretation:** Speech can support automatic assessment of schizophrenia symptoms using pretrained embeddings, even without transcripts. Such models show promise as clinically meaningful, efficient, and low-burden tools for real-time monitoring of symptom trajectories. Future work should validate these models prospectively in higher-severity samples and improve clinical interpretability.

**Funding:** European Union Horizon.

**Research in context:** *Evidence before this study:* Automatic assessment of disease severity is a key issue in schizophrenia research, for which spontaneous speech offers a cost-effective, automatable solution. To evaluate existing evidence for speech-based symptom assessment, two reviewers (RHe, MK) searched PubMed, IEEE Xplore, arXiv, bioRxiv, and medRxiv for publications from inception to Aug 25, 2025, using the terms: (“symptom” OR “PANSS” OR “Positive and Negative Syndrome Scale”) AND (“psychosis” OR “schizophrenia”) AND (“language” OR “speech” OR “spontaneous speech”) AND (“prediction” OR “machine learning” OR “deep learning” OR “algorithm” OR “neural network” OR “AI” OR “artificial intelligence”). Fourteen studies on symptom-level modelling were identified. Ten studies dichotomized clinical scores (e.g., Positive and Negative Syndrome Scale (PANSS)) into low vs high for classification: five used conventional machine learning (e.g., random forests) and five used neural networks, with F1 scores ranging from 0·60-0·85. The remaining four studies, and two of the ten studies as mentioned above, modelled raw scores directly as regression tasks. Two relied solely on conventional regressors and the rest used neural networks, with errors from 0·487 for single items (scale 1-7) to 8·04 for summed scores (scale 18-126). All studies used free speech for elicitation, except one study, which used a reading task. Three studies incorporated additional tasks, such as picture description and immediate recall. None were multilingual: nine were in English, three in Chinese, one in Swiss German, and one in Brazilian Portuguese. Features spanned a wide range, including acoustic-prosodic profiles, morpho-syntactic structure, semantic organization, pragmatics (including sentiments), and even visual features capturing movement during talking. Representations from pretrained language models were also widely employed. Sample sizes (counting patients with schizophrenia) were generally small: eleven studies enrolled <50 patients, one had 65, and only two exceeded 100 patients. Some increased their effective sample size via multiple recordings per patient or by adding healthy controls and/or patients with other psychiatric disorders (e.g., depression).

*Added value of this study:* In this computational secondary analysis of previously collected data, we aggregated speech recordings and clinical assessments across ten international sites. To our knowledge, this is the first multilingual, speech-based study modelling schizophrenia symptom severity with machine learning approach, and it includes the largest cohort of patients with schizophrenia to date. We further increased effective sample size by using diverse elicitation tasks and segmenting recordings into clips. This multilingual corpus empowers the usage of complex models and supports transfer learning from high-resource languages (e.g., English) with ample available speech and language computational resources to low-resource ones lacking these resources (e.g., Czech). For each of eight selected relapse-related PANSS items, the best audio-only models achieved root-mean-squared error (RMSE) < 1·5, underscoring clinical relevance. We assessed potential biases: no effects were found for age, sex, or education (except poorer N4 performance in males), though performance declined at higher symptom severity. Trained models are released for use.

*Implications of all the available evidence:* We show that speech is a powerful signal for automatic assessment of schizophrenia symptom severity and holds promise for relapse prediction, even without transcripts. The approach readily extends to incorporate textual features (from manual or automatic transcripts) and more advanced models. Prospective studies with repeated recordings across relapse episodes are needed to validate the utility of our models on relapse prediction, for the sake of supporting precision psychiatry while reducing clinician burden.

## Introduction

Symptoms in schizophrenia spectrum disorders / psychotic disorders fluctuate over time, often with remission phases and relapses.^1^ Relapse is typically operationalized as a clinically meaningful exacerbation of symptoms after a period of stability.^2^ Identifying relapse-predictive symptom fluctuations is a crucial desideratum but remains time-consuming and labour-intensive for both patients and clinicians. There is a clear need for cost-effective, scalable automation to streamline care, enable more frequent monitoring, and reduce clinician workload. Language, especially spontaneous speech, fits this remit. Conceptually, speech and language provide a readout of mental state dynamics that vary during relapse, and offers analytic and mechanistic substrates as the L-factor of the psychopathlogy.^3^ Language dysfunction in schizophrenia spectrum disorders (SSD) is well documented.^4,5^ It is linked to symptomatology^6,7^ and signals relapse risk.^8^ In addition, from a practical perspective, speech can be collected passively, remotely, and cheap, and analysed automatically at low marginal cost.

Efforts to automatically model language alterations in SSD have been made over decades thanks to the advances in speech and language techniques. Spontaneous speech consistently shows strong potential for identifying SSD,^9–15^ alerting psychotic risk,^16^ and capturing symptom severity,^17–22^ across disease stages, different languages, and elicitation tasks. Of these, modelling symptom severity is especially compelling given its connection to relapse prediction. The Positive and Negative Syndrome Scale (PANSS) provides an operationalizable, standardized outcome measure on 30 different symptoms across positive, negative, and general psychopathology domains, with each item scored from 1 to 7, where higher scores indicate greater severity. Although PANSS total score is indicative of relapse (when score increases surpass a threshold) and detectable from spontaneous speech,^17–20^ the full scale is often not used uniformly across clinical sites, with some centres rating only selected subscales. Accordingly, the present study focuses on eight symptoms that were most consistently assessed across sites and were specified in the study protocol as relapse markers,^23,24^ namely: three positive (P1. Delusions; P2. Conceptual Disorganization; P3. Hallucinatory Behaviour), three negative (N1. Blunted Affect; N4. Passive/Apathetic Social Withdrawal; N6. Lack of Spontaneity and Flow of Conversation), and two general items (G5. Mannerisms and Posturing; G9. Unusual Thought Content). Previous work has tried to dichotomize item scores into low vs high for classification, with accuracy of 0·80 on P2,^25^ and accuracies around 0·65^19^ and 0·85^11^ on negative items (especially blunted affect). However, previous work has paid relatively less attention to other relevant positive and general psychopathology items.

Key gaps persist despite decades of work. First, previous studies rely almost exclusively on monolingual data, predominantly English. Given empirical and meta-analytic evidence that voice-pattern alterations in schizophrenia vary across languages^26^, such an English-centric base limits cross-lingual generalisability. Low-resource languages, where speech datasets and computational or clinical linguistic resources remain limited, are often overlooked in schizophrenia research. This imbalance increases the risk of language-specific artefacts and reduces generalisability to real-world clinical and remote settings. Single-source datasets also embed collection biases from the recording chain and context (e.g. device, sampling, room noises, speaker-mic distance, elicitation tasks, interviewer’s questions, accents and dialects), risking confounds and undermining robustness. Second, most existing pipelines are transcript-dependent: speech must first be converted to text, either by manual transcription or automatic speech recognition (ASR), before linguistic features can be extracted. Manual transcription is costly and difficult to scale, while ASR performance degrades in noisy clinical recordings and for under-represented languages and accents, introducing additional errors and bias. In contrast, transcript-free, speech-only approaches operate directly on the audio signal and derived acoustic-prosodic features, avoiding the transcription costs. Third, available datasets are typically small, often including fewer than 50 individuals with SSD, which limits model performance, external validity, and scalability. Progress therefore hinges on training predictive models in larger, multicentre, multilingual samples using representations relying only on speech, so that information contained in speech can be exploited without relying on transcripts or language-specific resources.

Therefore, in this study, we pooled speech data from ten centres across multiple languages (or variants of the same language) and from patients in different stages of disease, yielding the largest SSD speech corpus to date by patient count. Recordings were processed without transcripts to extract acoustic-prosodic profiles and pretrained multilingual speech embeddings, from which predictors for eight PANSS items were trained with tuned hyperparameters to enhance performance and generalisability. In addition, cross-language and cross-task transfer were evaluated, and potential demographic biases from the models were systematically assessed. Finally, an independent dataset was introduced to probe model generalisability. Overall, we indicate that speech carries rich signal for symptom-severity assessment and generalizes across languages, disease stages, and collection settings, supporting its use for relapse-risk stratification and the integration into routine psychiatric care.

## Methods

### Study design and participants

This study was a secondary analysis of de-identified speech recordings and item-level PANSS ratings collected across multiple international sites. The workflow of the study is indicated in **Figure 1**. Data from a total of 453 patients with SSD was included, all recruited from different primary studies across ten international centres across different languages between 2005 and 2026: Czech Republic (Czech, cs, *n* = 16), the United States (English, en, *n* = 42), Spain (Spanish, es, *n* = 48), Chile (Chilean-Spanish, escl, *n* = 14), France (French, fr, *n* = 16), Switzerland (Swiss German, gsw, *n* = 35), Netherlands (Dutch, nl, *n* = 88), Türkiye (Turkish, tr, *n* = 108), Germany (German, de, *n* = 40), and China (Chinese Mandarin, zh, *n* = 46). Diagnoses were made by local psychiatrists using applicable diagnostic criteria. Participants with missing symptom scores or with poor-quality, missing or corrupted audio recordings were excluded. Age, sex, and years of education are reported in **Table 1** for every site, and their distributions were visualized in **Supplementary Figure 1-4**. Data on participant race and ethnicity were not explicitly collected in this study. The dataset was aggregated from a multi-national consortium spanning over a dozen countries. Not all sites have collected such information and some sites were not allowed to collect them. However, the broad geographic and linguistic representation, encompassing diverse populations across European, Asian, and American countries, mitigates the risk of findings being limited to a single cultural or linguistic context. Data used in this secondary analysis were derived from pre-existing research studies and cohorts conducted at the participating centres rather than collected specifically for the present study. The source study or cohort, data-contributing institution, and relevant publications are detailed in the supplementary. For each centre, the original data were accessed and contributed by the local investigators, all of whom are co-authors of the present study and were authorised under the applicable ethical approvals and institutional data-governance arrangements to use and contribute the corresponding data for this secondary analysis.

**Figure 1.**
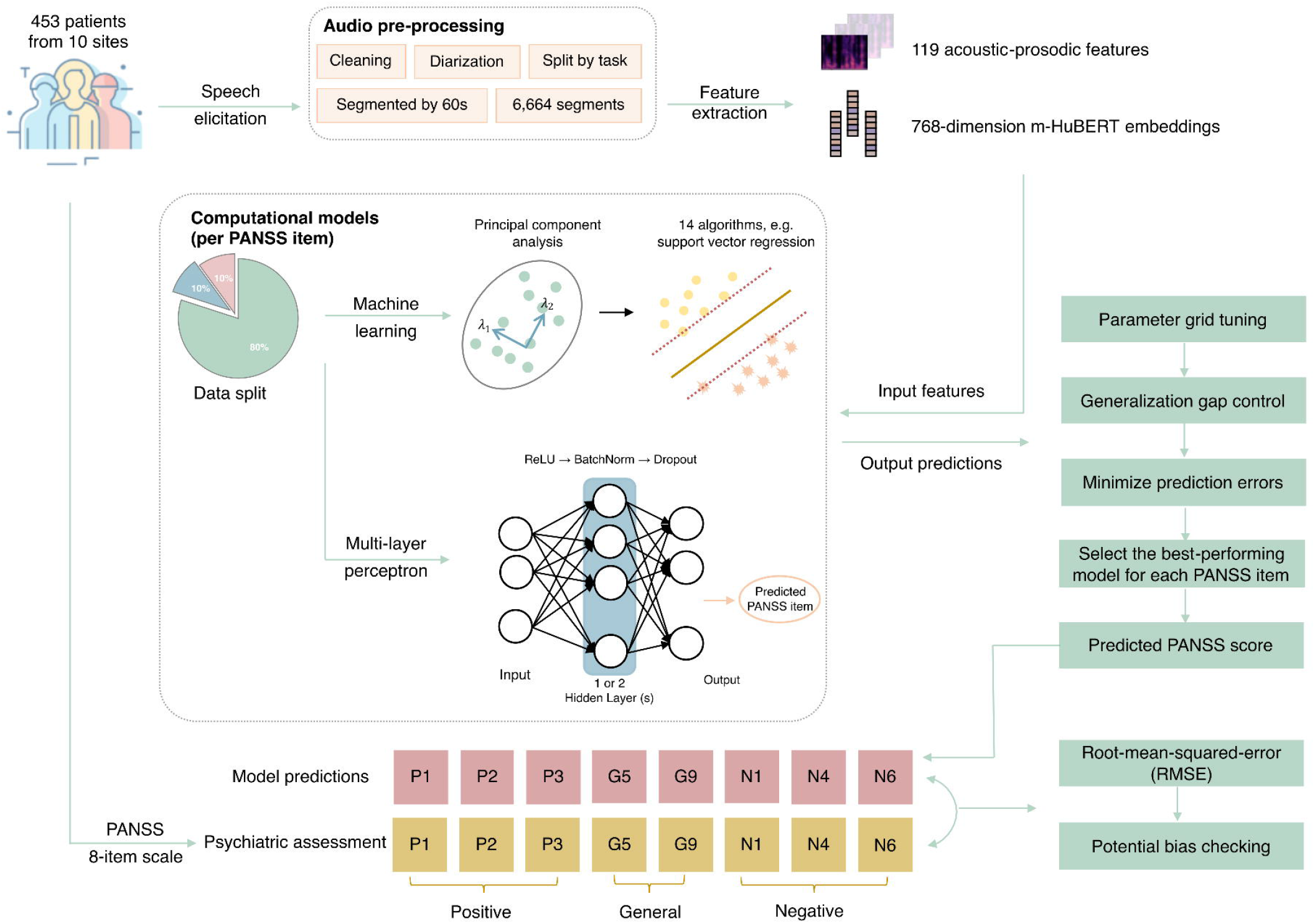
Workflow of this study. Speech recordings from 453 patients across 10 sites were cleaned, diarized, split by task, and segmented into 60-s windows, yielding 6,664 speech segments. From each segment, 119 acoustic-prosodic features and 768-dimensional m-HuBERT embeddings were extracted as model inputs. The data were split into training, validation, and test sets, and computational models were trained separately for each target PANSS symptom score. The MLP schematic illustrates one symptom-specific single-output model, producing one predicted PANSS score for a given target symptom rather than simultaneously predicting all eight symptoms. Model development included parameter grid tuning, generalization-gap control, and validation-error minimization. The best-performing model was selected separately for each PANSS symptom, and final predictions were evaluated against clinician-rated PANSS scores using root-mean-squared error (RMSE) and potential prediction bias checks. PANSS items: P1. Delusions; P2. Conceptual Disorganization; P3. Hallucinatory Behaviour; N1. Blunted Affect; N4. Passive/Apathetic Social Withdrawal; N6. Lack of Spontaneity and Flow of Conversation; G5. Mannerisms and Posturing; G9. Unusual Thought Content. PANSS = Positive and Negative Syndrome Scale; MLP = multi-layer perceptron; RMSE = root-mean-squared error; m-HuBERT = multilingual HuBERT; ReLU = rectified linear unit; BatchNorm = batch normalization.

### Ethics

All procedures followed local ethical approvals, including: the ethics commission of National Institute of Mental Health (NIMH) of the Czech Republic; IRB at Feinstein Institutes for Medical Research, Northwell Health; local institutional board at Valdecilla Research Institute (IDIVAL) in Santander, Spain; Ethics Committee for Clinical Research, CEC SSMS of Santiago, Chile; ethical commission of Montperrin Hospital, CH Aix-en-Provence, France; the ethics committee of Kantonale Ethikkommission Zurich; review boards of the University medical center Utrecht (for RAPSODI trial) and Groningen (for HAMLETT cohort); the Ethics Committee of Dokuz Eylul University; Ethics Committee of the State Chamber of Physicians Westphalia-Lippe and the University of Muenster; the ethics committee of Renmin Hospital of Wuhan University and the Institutional Review Board of the Institute of Psychology, the Chinese Academy of Sciences; and the Clinical Research Ethics Committee of the Hospital de la Santa Creu i Sant Pau. Written informed consent was obtained from all participants. The studies are adherence to the Declaration of Helsinki. Details on the recruitment, inclusion and exclusion criteria, and ethical approval are available in the supplementary (p 2-4).

### Procedures

All participants were assessed for clinical symptoms. Most had ratings on at least 8 items^24^ of the PANSS scale, including three positive (P1, P2, P3), three negative (N1, N4, N6), and two general (G5, G9) items. These items, as mentioned above, are considered as markers of relapse in SSD. All PANSS items were rated on a 7-point scale from 1 to 7, where 1 indicates least symptom load and 7 indicates greatest symptom load. PANSS was not administered at some sites: the English-speaking participants were assessed with the Brief Psychiatric Rating Scale (BPRS) and Scale for the Assessment of Negative Symptoms (SANS), while Turkish-speaking and some Chinese-speaking participants were assessed with the Scale for the Assessment of Positive Symptoms (SAPS) instead. Their scores were converted to corresponding PANSS item scores using previously published formulas.^27^ Clinical scale assessments were conducted locally within the original studies using instruments provided and maintained by the participating hospitals or clinical institutions. The present retrospective study received and analysed only de-identified item-level scores and did not obtain, reproduce, or redistribute the scale materials. **Table 1** summarizes the group-level scores of each item and global scores. We modelled and evaluated each of these eight PANSS items. All participants completed at least one spontaneous speech task. Speech protocols varied across sites, but overall, the tasks fell into five categories: (1) free speech (conversational or interview-style narration), (2) picture description (structured description of visual scenes), (3) dream report (narration of remembered dreams), (4) reading (oral reading of given texts), and (5) recall (retelling of short stories or video clips immediately). Details and task classification are provided in the supplementary (p 4).

### Audio processing and feature extraction

Recordings were manually diarized by trained annotators to remove the interviewer’s voice, segmented by task, and further divided into intervals of ≤60 seconds to harmonize length and facilitate downstream applications. Recordings shorter than one minute were left unsegmented. All segments derived from the same recording inherited the same PANSS item score. This procedure was applied automatically, resulting in 6,664 segments (**Table 1**). We first extracted acoustic-prosodic features. These features were automatically extracted using OpenSmile (eGeMAPSv02, at the level of functionals), supplemented with Prosogram for finer-grained prosodic analysis. Together, these descriptors capture multiple aspects of speech wave dynamics, including pitch variation, loudness, speech rate, pausing behaviour, spectral balance, cepstral properties, temporal organization, and voice quality. These voice patterns have been reported as markers of schizophrenia with a certain level of cross-lingual generalisability.^26^ In addition, recordings were encoded using utter-project/mHuBERT-147, a multilingual speech representation model. This model converts the waveform into high-dimensional embeddings that capture the joint patterns of articulation, prosody, and segmental context that may not be fully represented by predefined acoustic-prosodic descriptors alone. In this sense, the embeddings provide a richer and less manually constrained representation of the speech signal, capturing higher-order regularities related not only to low-level acoustics, but pronunciation patterns and rhythmic structure, and even syntactic structure and conceptual knowledge as encoded in the audios.^28^ Together, these features were chosen to combine clinically motivated handcrafted speech measures with transcript-free multilingual representations, in line with the study’s aim of developing a scalable and practically deployable speech-based marker across sites and languages. All tools are publicly available and scalable across populations without necessity for manual annotation. (supplementary p 4)

### Computational modelling

Segments were split into training (80%), validation (10%), and test (10%) sets by participant within each language, stratifying by a binary severity label (at least two items over three vs. not)^23^ to preserve severity and language distributions and prevent leakage (number and ratio available in **Supplementary Table 1**). We first evaluated 14 machine-learning algorithms on three feature sets (acoustic-prosodic, m-HuBERT, concatenation). Algorithms included ordinary least squares, ridge, Bayesian ridge, lasso, elastic net, Huber, support vector regression, random forest, extra trees, gradient boosting, AdaBoost, XGBoost, k-nearest neighbours, and Gaussian process regression. Features were standardized and reduced by principal component analysis (PCA). We then trained two feedforward neural networks (Multi-Layer Perceptrons, MLPs), with two and three layers, respectively. Each hidden layer applied nonlinear activation followed by batch normalization. Training used AdamW with RMSE loss for up to 100 epochs, batch size 128. Early stopping was applied if validation performance did not improve over 0·01 for four consecutive epochs. To mitigate overfitting, we applied dropout after batch normalization in each hidden layer, and adjusted learning rate using the OneCycleLR scheduler with L2 regularization. Details are provided in the supplementary (p 5).

### Hyperparameter tuning

For each PANSS item, we trained the models and applied grid search over candidate hyperparameter configurations to tune the hyperparameters. Models were trained on the training set and evaluated on the validation set with RMSE, with lower values indicating better predictive accuracy. For each PANSS item, candidate configurations were first filtered by their generalization gap on the validation data to reduce overfitting risk. The generalization gap was defined as the relative difference between validation and training RMSE. Only models with positive gap under 0·5 were retained. Within each model-feature combination, the configuration with lowest validation RMSE was retained. Secondly, these configurations were compared on the test set, and the final model for each PANSS item was chosen as the one with the lowest test RMSE and acceptable test generalization gap. Details are provided in the supplementary (p 5-6).

### Performance evaluation

For each PANSS item, the final selected model was evaluated on the held-out test set. We reported RMSE as the primary metric. We additionally reported R-squared as a goodness-of-fit measure in the supplementary materials. Performance was assessed at two levels, of segments and of participants. Metrics were first reported at the segment level for the entire dataset as well as separately for each language. To account for variability across segments, predictions were further aggregated at the participant level using the median score across segments, and corresponding performance of RMSE was reported. Details are provided in the supplementary (p 6-7), with learning curves as in **Supplementary Figure 5-12**. As a supplementary analysis, participant-level aggregation based on averaging was also reported in **Supplementary Table 2**.

### Statistical comparisons and potential bias checking

Within each model-feature combination, the configuration with the lowest validation RMSE was retained. To compare predictive performance across symptom targets and feature sets, we used the aligned rank transform (ART), a nonparametric factorial procedure that enables the estimation of main effects and interactions while relaxing the normality assumptions required by conventional analysis of variance (ANOVA). In this analysis, test-set RMSE served as the dependent variable, symptom and feature were entered as fixed factors, and model was included as a blocking factor to account for repeated comparisons across model architectures. ANOVA-style tests were then conducted on the aligned and ranked data to evaluate the main effects of symptom and feature, as well as their interaction. When significant omnibus effects were observed, post-hoc pairwise comparisons were performed and corrected for multiple testing using the false discovery rate (FDR) procedure.

To check potential bias in PANSS prediction from basic demographics, elicitation tasks, and symptom severity, we focused on the best-performing models and summarized predictions first at the segment level and then at the participant level. At the participant level, absolute prediction errors were examined for potential demographic and clinical biases. Specifically, we assessed associations between absolute prediction error and age, years of education, and symptom severity (i.e., the observed PANSS item score) using correlation analyses. To test whether prediction error differed by sex, we used permutation testing, which provides a distribution-free assessment of group differences and is suitable for the present sample size and data structure. To test whether model accuracy differed systematically by elicitation condition, segment-level predictions belonging to the same task were aggregated using the median in order to reduce the influence of extreme values and local variability across segments. Absolute prediction error, defined as the absolute difference between predicted and observed PANSS item scores, was then compared across tasks using ART. Statistical significance was defined as a two-sided *p* value below 0·05, with FDR correction applied where multiple comparisons were conducted.

### Out-of-sample deployment

To test the utility of the model on out-of-sample data, we additionally analysed a pilot dataset from an ongoing local study. Five participants completed a baseline full speech protocol and 30-item PANSS. Around three months later, they returned for a follow-up with a subset of speech tasks (mainly picture descriptions) and a reduced PANSS (three positive, and three negative items). This dataset differs from our held-out test set in three ways: (i) its distribution was not matched to the training data; (ii) two participants spoke Spanish (present in training) while three spoke Catalan (absent from training); and (iii) participants were substantially older. We used it to probe external generalisability, applying the best-performing models and reporting absolute errors. Age, sex, years of education, and PANSS scores are reported in **Table 2**. Details are provided in the supplementary (p 7).

### Prespecified and post-hoc analyses

All primary outcomes and analyses related to model development, performance evaluation, statistical comparison across models and feature sets, and potential bias checking were prespecified before model training and evaluation. No additional post-hoc analyses were added to the main Results during peer review. The out-of-sample deployment analysis was not part of the original model-development plan. It was added after model development when a small independent dataset became available, and was therefore labelled as an exploratory post-hoc analysis.

### Software

All analyses were performed mainly using self-developed Python (version 3.11.14) and R (version 4.5.2) scripts. Feature extraction mainly utilised opensmile (version 2.6.0), librosa (version 0.11.0), pydub (version 0.25.1), transformers (version 4.28.0) and the Prosogram praat toolkit (version 3.01b). Model training, testing, and validation mainly utilised sklearn (version 1.7.2), xgboost (version 3.2.0), and torch (version 2.0.0). Statistical analyses and visualization mainly utilized R packages, including coin (version 1.4.3), readr (version 2.1.5), tidyr (version 1.3.1), dplyr (version 1.1.4), purrr (version 1.2.2), stringr (version 1.5.2), ggplot2 (version 4.0.0), ARTool (version 0.11.2), readxl (version 1.4.5), emmeans (version 2.0.3), multcomp (version 1.4.28), and ggpubr (version 0.6.1).

### Role of the funding source

The funder of the study had no role in study design, data collection, data analysis, data interpretation, or writing of the report.

## Results

### Model performance

Figure 2. summarized the model performance. We first identified the optimal parameter set for each model-feature combination, as shown in **Figure 2**A. In **Figure 2**B and C, we reported whether model performance was comparable across PANSS items and speech feature sets. ART analyses indicated that prediction error was lowest for G5 and N6, followed by N1 and G9, then P2 and P3, and highest for P1 and N4 (**Figure 2**C). Models using m-HuBERT embeddings outperformed those with concatenated features, which in turn surpassed models based solely on acoustic-prosodic features (**Figure 2B**). All pairwise differences and item-feature interactions were statistically significant. We then identified a single best-performing model for each item. Across items, m-HuBERT embeddings consistently supported the best predictions, except for N4 and G5, where models using acoustic-prosodic features yielded the lowest errors. A support vector regressor best predicted P2 (segment: RMSE=1·318, R^2^=0·280; participant: RMSE=1·107, R^2^=0·321). A two-layer MLP best predicted P1 (segment: RMSE = 1·544, R^2^ = 0·237; participant: RMSE = 1·527, R^2^ = 0·176), N4 (segment: RMSE = 1·521, R^2^ = 0·093; participant: RMSE = 1·430, R^2^ = 0·183), and G5 (segment: RMSE = 0·951, R^2^ = −0·021; participant: RMSE = 0·882, R^2^ = 0·070). A three-layer MLP best predicted P3 (segment: RMSE = 1·487, R^2^ = 0·195; participant: RMSE = 1·542, R^2^ = 0·138), N1 (segment: RMSE = 1·071, R^2^ = 0·269; participant: RMSE = 1·030, R^2^ = 0·280), N6 (segment: RMSE = 0·869, R^2^ = 0·120; participant: RMSE = 0·855, R^2^ = 0·209), and G9 (segment: RMSE = 1·298, R^2^ = 0·429; participant: RMSE = 1·282, R^2^ = 0·376). Results for each language are displayed in **Figure 2**D and E and **Supplementary Tables 3 and 4.**Performance at the segment and participant levels were relatively consistent. Specifically, the predictive error for French and Chinese was consistently higher (higher RMSE) across all symptom targets, whereas Peninsular and Chilean Spanish, and German and Dutch yielded more favourable outcomes (lower RMSE). The specific parameters of the best models are described in the supplementary (p 8).

**Figure 2.**
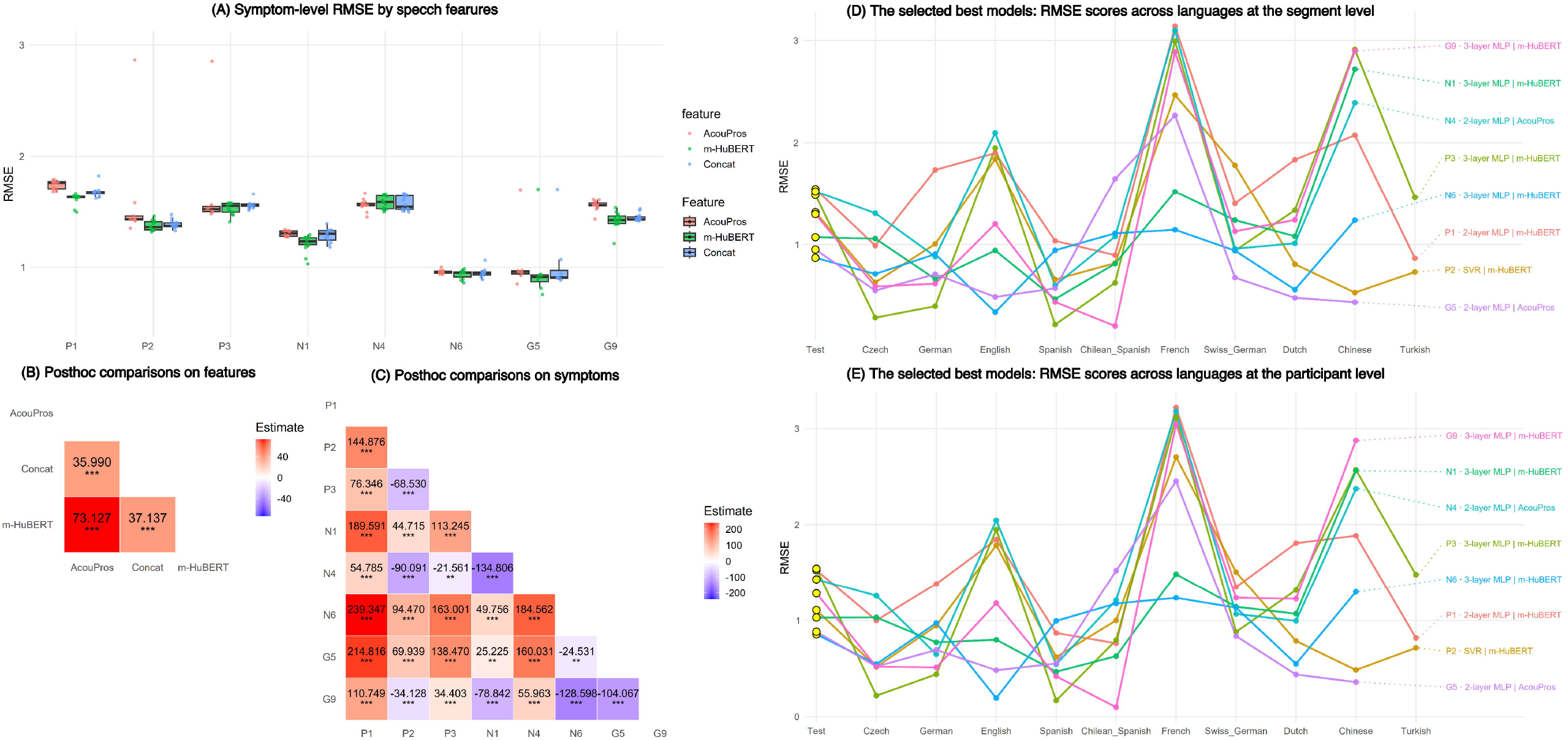
Summary of model performance. (A) Prediction errors (RMSE) for each item across feature sets (acoustic-prosodic (AcouPros), m-HuBERT, and concatenation of both two). (B) Pairwise post-hoc comparisons across feature sets to compare model performance with different features, with effect size estimates and significance levels. *** p < 0·001; ** p < 0·01; * p < 0·05. Red values indicate that the feature set in the row had significantly better performance than the feature set in the column, while purple values indicate significantly worse performance. (C) Pairwise post-hoc comparisons across items to compare model performance on different items. *** p < 0·001; ** p < 0·01; * p < 0·05. Red values indicate that the feature set in the row had significantly better performance than the feature set in the column, while purple values indicate significantly worse performance. (D) Best-performing models for each item, with RMSE scores across languages, at the segment level. Coloured labels on the right indicate the corresponding models and features. (E) Same as (D), but evaluated at the participant level. PANSS items: P1. Delusions; P2. Conceptual Disorganization; P3. Hallucinatory Behaviour; N1. Blunted Affect; N4. Passive/Apathetic Social Withdrawal; N6. Lack of Spontaneity and Flow of Conversation; G5. Mannerisms and Posturing; G9. Unusual Thought Content. RMSE = root-mean-squared error; PANSS = Positive and Negative Syndrome Scale; AcouPros = acoustic-prosodic features; m-HuBERT = multilingual HuBERT; MLP = multi-layer perceptron; SVR = support vector regression; Concat = concatenated acoustic-prosodic and m-HuBERT features.

### Bias checking

Figure 3. summarized the potential bias. As shown in **Figure 3**A–C, prediction errors were not significantly associated with age, sex, or education in most analyses. The only exception was the MLP model for N4, which showed significantly larger prediction errors in male participants than in female participants (**Figure 3**B, N4: FDR-corrected *p*=0·042, other items with FDR-corrected *p*>0·05). Overall, these results provide little evidence of systematic demographic bias, although larger samples will be needed to assess such effects more conclusively. Likewise, ART analyses revealed no significant differences in prediction errors across elicitation tasks (**Figure 3**E), suggesting that model performance was broadly comparable across subtasks. In contrast, correlational analyses suggested significant patterns with symptom severity. Prediction errors were significantly associated with symptom severity for all items except N4 (**Figure 3**D, FDR-corrected *p*>0·05), with higher observed symptom scores generally linked to larger errors. This pattern suggests that more severe symptom presentations were harder for the models to predict accurately.

### Performance on out-of-sample data

The models performed relatively, and surprisingly, well on the external validation data. The RMSE scores were comparatively low in all items, except P3 (P1: 1·640, P2: 0·666, P3: 3·192; N1: 1·183, N4: 1·305, N6: 1·017; G5: 0·352, G9: 1·029), with details reported in **Table 2**.

**Figure 3.**
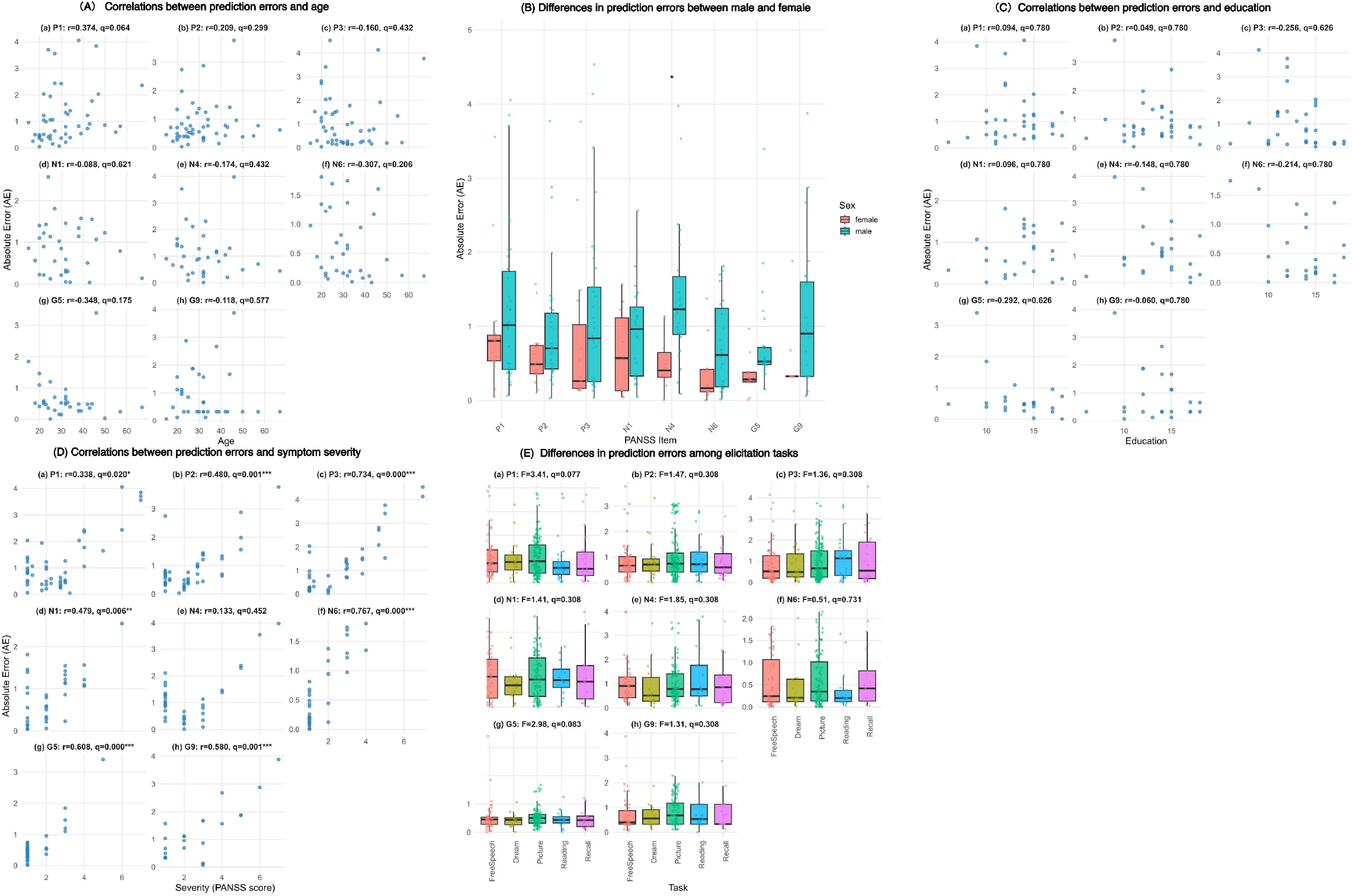
Results of bias checking. Boxplots show the median, interquartile range, and whiskers extending to 1·5 times the interquartile range. Dots represent individual observations. *q* values denote false-discovery-rate-adjusted *p* values. (A) Scatter plots of prediction errors (absolute error, AE) against age for each PANSS item: (a) P1, Delusion; (b) P2, Conceptual disorganization; (c) P3, Hallucinatory behaviour; (d) N1, Blunted affect; (e) N4, Passive/apathetic social withdrawal; (f) N6, Lack of spontaneity and flow of conversation; (g) G5, Mannerisms and posturing; and (h) G9, Unusual thought content. Subtitles indicated the correlation coefficients (*r*) and adjusted *p* values (reported as *q*). (B) Boxplots with strip plot overlays of prediction errors by sex for each PANSS item. Comparisons with asterisks were significant. *** p < 0·001; ** p < 0·01; * p < 0·05. (C) Scatter plots of prediction errors against education, with same settings as (A). (D) Scatter plots of prediction errors against symptom severity, with same settings as (A). Correlations with asterisks were significant. *** p < 0·001; ** p < 0·01; * p < 0·05. (E) Boxplots with strip plot overlays of prediction errors by task categories. AE = absolute error; PANSS = Positive and Negative Syndrome Scale.

## Discussion

The present study, to our knowledge, reflects the largest effort so far to predict individual PANSS item scores in a diverse, multi-site and multilingual setting using only audio recordings from several speech elicitation tasks. Our analysis covers both lower-(e.g. Czech) and higher-resource languages (e.g. English, Chinese), as well as variants of the same language (e.g. peninsular and Chilean Spanish). For 8 PANSS items selected for their clinical relevance to relapse, we show that, in the absence of transcriptions, RMSE scores can reach 1·5 or lower, indicating practically useful accuracy at the item level. The best-performing model relied on self-supervised speech embeddings extracted directly from the audio, without hand-crafted acoustic or textual features.

Few prior studies are directly comparable, because existing work differs substantially in target scale (e.g., PANSS, Brief Psychiatric Rating Scale (BPRS), Negative Symptom Assessment-16 (NSA-16), or Thought, Language and Communication (TLC) scale), prediction granularity (item-level vs. subscale vs. overall severity), speech elicitation task, and cohort composition,^17–20^ or used a classification approach dichotomizing scores into high vs low.^11,19,25^ Voleti et al. combined features from multiple linguistic domains to regress on the averaged PANSS positive and negative scores, to get mean squared error around 0·5.^17^ Rohanian et al. fused speech with textual features in gated recurrent neural network to predict the total positive and negative scores with RMSE around 3·5 to 4.^18^ Chuang et al. constructed transformers with fusion networks to utilize multimodal information from speech, text, and vision and reported PANSS item-level mean squared error of 1·111.^20^ All of these studies were conducted within a single cohort and focused on Indo-European languages. In contrast, our models used speech alone, without relying on manual transcription, linguistic annotation, or other labour-intensive preprocessing steps. Despite this substantially leaner and more scalable pipeline, our models achieved performance comparable to, and in some cases better than, prior work. This is particularly notable because our models were evaluated in a cross-linguistic setting and in a larger, more heterogeneous, and clinically variable cohort, all of which are likely to increase task difficulty. At the same time, this design broadens the potential applicability of the approach by enabling the inclusion of previously underrepresented languages and populations in model development.

Of note, a traditional assumption, reflected in previous work,^11^ has targeted negative symptoms as those predictable from acoustic-prosodic features. Given the high cost of transcription and the fact that transcription was not permitted for part of the dataset, we did not include transcript-derived features in the present study, and instead reserve text-based PANSS prediction for future research. Our study reveals that acceptable error rates extend to positive and general items of the PANSS. This is consistent with recent model-interpretability work using layer-wise minimal-pair probing of self-supervised speech models, showing that although these models are trained only on raw audio, their representations capture both syntactic constraints and semantic contrasts beyond low-level acoustics. Such embeddings may therefore carry linguistically rich information that is relevant to a broader range of symptom dimensions.^28^ At the same time, a crucial nuance in our findings is that performance appears to degrade with higher symptom severity: models are systematically less accurate at the upper end of the score range (**Figure 2**D).

This may arise from the fact that PANSS score distributions in our dataset were imbalanced. Most observations were concentrated at lower symptom levels, whereas the highest scores were rare (**Supplementary Figure 13**). We note that this issue was considered during data splitting: the train, validation, and test sets were stratified according to whether the patients had at least two PANSS scores above 3. However, such stratification can only distribute the available high-severity cases across subsets, and it cannot compensate for severity levels that are extremely rare in the dataset. This imbalance may reflect a common feasibility-related restriction in clinical speech datasets: patients with very severe symptoms are often more difficult to recruit for speech-based studies. They may be less able to provide informed consent, may have difficulty completing structured speech elicitation tasks, or may produce insufficient analysable speech, particularly in the presence of severe negative symptoms. Thus, the under-representation of the highest PANSS levels reflects a broader practical challenge in collecting speech data from the most clinically severe cases. Nevertheless, this distributional restriction has important consequences for model interpretation. It was also reflected in the prediction scatter plots (**Supplementary Figure 14**), where models showed restricted prediction ranges and a systematic tendency to under-predict the upper end of the severity scale. Accordingly, the validity of the present models is currently best supported for the lower-to-moderate symptom range represented in the dataset. For higher severity levels, especially when empirical examples were absent from training, model outputs would necessarily involve extrapolation beyond the observed data distribution. Of note, the **Supplementary Figures 13 and 14**, with the underlying analysis, were added post hoc during the peer review.

Note that, as for clinical applicability, from the perspective of relapse prediction, reduced precision at very high symptom levels may be less critical, since marked exacerbations are usually clinically obvious, whereas small shifts in the lower-to-moderate range, where our models perform better, may provide earlier, and therefore more actionable, warning signals for impending relapse. However, this potential application does not remove the need to validate speech-based models in high-severity cases before they can be considered reliable across the full clinical spectrum. Future work should therefore address this limitation by including cases with higher severity and via data augmentation.

We observed no systematic evidence for large demographic or task-related biases. Bias checks suggested no major effects of age, sex, education, or task on prediction errors overall, with the exception of N4, where predictions for male participants were worse. This pattern cautions against strong claims of complete demographic neutrality, but it does support the view that a speech marker based on self-supervised representations can be made reasonably scalable and low-cost across different subgroups if subgroup performance is continuously monitored. Task-insensitivity of error rates is noteworthy, as prior studies using transcription-based features to predict diagnostic groups have shown task sensitivity.^29^ A digital speech marker based on embeddings of audios only, irrespective of task, may be highly scalable across demographically diverse samples. Moreover, speech-based acoustic and prosodic features have recently been shown to be satisfactorily stable across measurement points in neurotypical individuals,^30^ unlike text-based features, recommending such features in psychometric terms for clinical applications as well.

Results of this study should be considered with several limitations. This study used a heterogeneous dataset in terms of samples, languages, symptoms, symptom assessment, and speech elicitations. This heterogeneity was partially intentional, as it reflects the heterogeneity of real-world naturalistic data, against which predictive models need to be robust. However, it limits exploration of additional questions of interest, such as which language-specific differences affect predictions. In our setting, differences between languages across sites are confounded by differences across sites, along a number of other dimensions, such as symptom distributions and speech elicitations. Here, our primary focus was on generalisability of the model across samples and languages, rather than investigating language-specific patterns. Additionally, despite the fact that bias check suggested no evidence for bias on age, sex, and education, our results do not eliminate a potential concern of a racial and ethnic biases when using AI-based language models. Second, what we describe as an out-of-sample deployment in a separate cohort of five individuals should be viewed as a small real-world use case to preliminarily probe generalisability. Larger and more diverse deployment samples will be needed to draw strong conclusions about external validity.

Another important limitation concerns the PANSS ratings themselves. Scale-based clinical ratings, and the PANSS in particular, show substantial inter-rater variability across sites. In this multi-centre, retrospective dataset we could neither standardize assessment procedures nor obtain centralized ratings, so the targets that our models learn from are inevitably noisy. This level of noise in the data likely depresses attainable performance and may blur some item-specific effects, but it also means that the models are optimized to make predictions that are robust to the kind of variability encountered in real-world rating practice, rather than representing idealized noise-free scores.

Finally, our predictors remain largely black boxes. We did not undertake detailed model-explainability analyses in the present study because the primary aim was first to establish whether transcript-free symptom prediction was feasible in a deliberately difficult setting: speech alone, without manual transcription or linguistic annotation, across heterogeneous and cross-linguistic cohorts. Given the modest performance of some models and the imbalance at higher PANSS scores, we believe that detailed clinical interpretation of model features would be premature before predictive robustness and generalisability are more firmly established. Consequently, the current models should be viewed as preliminary decision-support candidates rather than clinically interpretable tools ready for deployment. Future work should therefore combine prospective validation with explicit explainability analyses. In the context of brain-encoding models that align language or speech embeddings with neural activity, there are opportunities to improve interpretability by examining how speech embeddings predict brain responses, thereby providing neurophysiological grounding. Another crucial avenue is to investigate the internal computational mechanisms of the models. Future work should include fine-tuning on more balanced and carefully curated datasets, explicitly modelling the full severity range, and exploring larger speech and language models that can leverage advances in natural language processing while remaining transparent and clinically interpretable.

## Contributors

RHe conceptualised the study, developed the methodology, wrote the software, performed validation and formal analyses, contributed to investigation and visualisation, and wrote the original draft. MK contributed to methodology, formal analysis, investigation, visualisation, and writing of the original draft. CP contributed to methodology, formal analysis, investigation, visualisation, and review and editing of the manuscript. LNO, SB, SiB, SC, AD, CD, RF, JTGM, GG, RHü, KK, AL, RQ, AS, VT, BV, HW, BY, SY, YZ, MA, RAA, EB, JdB, AFB, SK, MM, AP, FS, and SXT contributed resources and data curation. LP contributed resources, data curation, project administration, and review and editing of the manuscript. IES, PH, and WH conceptualised and supervised the study, acquired funding, administered the project, contributed resources and data curation, and contributed to writing of the original draft and review and editing of the manuscript. RHe and MK accessed and verified the underlying data. All authors had full access to the data relevant to their contributions, read and approved the final version of the manuscript. RHe, MK, and WH have accessed and verified the data. RHe, and WH were responsible for the decision to submit the manuscript.

## Data sharing

The original participant-level data, including speech recordings, transcripts, and clinical data, cannot be made publicly available because they were collected across multiple sites under site-specific ethical approvals, informed-consent procedures, and data-governance agreements that do not permit unrestricted redistribution. The associated individual-level data and data dictionary are therefore not deposited in a public repository. Enquiries regarding the data may be directed to the corresponding author. However, all scripts and results are available at https://github.com/RuiHe1999/PANSS_prediction. No additional study documents are publicly available through this repository.

## Declaration of interests

LP reports personal fees for serving as chief editor from the Canadian Medical Association Journals, speaker/consultant fee from Janssen Canada and Otsuka Canada, SPMM Course Limited, UK, Canadian Psychiatric Association; book royalties from Oxford University Press; investigator-initiated educational grants from Janssen Canada, Sunovion and Otsuka Canada outside the submitted work. IES reports charity grant from Janssen, speaker fee from Otzuka and Ludbeck. SXT owns equity and serves on the board for North Shore Therapeutics, received support for travelling and attending conference from ISCTM, is on the advisory board and owns equity for Psyrin, and serves as a consultant for Catholic Charities Neighborhood Services, Cogntiv, Inc., Wellcome Trust and LB Pharmaceuticals. PH has received grants and honoraria from Novartis, Lundbeck, Takeda, Mepha, Janssen, Boehringer Ingelheim, Neurolite and OM Pharma outside of this work. FS reports grants from the Ministry of Health of the Czech Republic and INTER-ACTION-LUAUS25 outside the submitted work, has a pending patent application (Application No. 19/228,402), and reports academic and research appointments at the Third Faculty of Medicine, Charles University, and the National Institute of Mental Health, Czech Republic. IES, JB, LNO, MK, SK, SC, WH, CP, and RHe report support for attending meetings and/or travel through European Research Council (ERC) and/or European Union Horizon Europe grant funds related to this work. RHe received travel support from the Schizophrenia International Research Society through an Early Career Award related to this work. All other authors reported no conflict of interests.

## Acknowledgments

We thank all participants, their family members, health-care professionals, leads, data managers, and research assistants for their support to this work. This work is part of the project, a TRUSTworthy speech-based AI monitorING system for the prediction of relapse in individuals with schizophrenia (TRUSTING), funded by the European Union’s Horizon Europe research and innovation programme under grant agreement No 101080251, to IS, PH, and WH. Views and opinions expressed are, however, those of the author(s) only and do not necessarily reflect those of the European Union or the European Health and Digital Executive Agency (HaDEA). Neither the European Union nor the granting authority can be held responsible for them. Authors are listed in an alphabetical order, except for local members of the leading research group and the TRUSTING PIs. Additional funders for data collection are as below. English data: NIMH/NIDCD K23 MH130750 and the Brain and Behavior Research Foundation Young Investigator Grant (to SXT). Spanish data: the Carlos III Health Institute (PI14/00639, PI14/00918, PI17/00221, PI20/00066 and PI23/00076, to RAA). Chilean Spanish data: the National Agency for Research and Development (ANID), Chile (Fondecyt Regular Grant No. 1241618, to AFB). Swiss German data: Swiss National Science Foundation (Grant No. 501100001711-191938, to PH), the Brain & Behaviour Research Foundation (Grant No. 28997, to PH), and the OPO Foundation (Grant No. 2020-0075, to PH). Dutch data: the RAPSODI study by ZonMW in the Netherlands (Grant No. 80-83600-98-40120, to IS), as part of the research program Rational Pharmacotherapy (Goed Gebruik Geneesmiddelen) (Grant No. 836041008, to IS), and HAMLETT study by ZonMW in the Netherlands (Grant No. 80-84800-98-41015, to IES). Turkish data: Scientific and Technological Research Council of Turkey (TUBITAK 2247 Project No: 120C141). Czech data: Ministry of Health of the Czech Republic (Grant No. NU22-04-00143, to FS). In addition, RAA was financed by a Miguel Servet contract from the Carlos III Health Institute (Grant No. CP18/00003) and a Consolidator Grant from the Ministerio de Ciencia e Invovación (Grant No. CNS2022-136110). AP was supported by a Marie Skłodowska-Curie Actions-H2020-MSCA-IF-2018 grant (ID: 832518, Project: MOVES). A.S was supported by the Carlsberg Foundation. KK was supported by Japan Society for the promotion of Science (JSPS) (PE, to AP). RHe was financed by the China Scholarship Council (Grant No. 202108390062) during part of this work, and is currently funded by the DELTA-Lang project (Synergy Grants 2023, Grant No. 101118756). The wide collaboration behind this work was made possible largely through the support of the DISCOURSE in Psychosis Consortium (https://discourseinpsychosis.org/), whose assistance was also instrumental in developing the speech assessment protocol for data collection at the separate sites.

## Tables

**Table 1.** Demographics, clinical assessment, and basic speech descriptors of the participants.

**Table 2.** Demographics, clinical assessment, and model performance of the external validation dataset.

## Notes

### Summary of Updates

Title, main text, and supplementary.

