## supplementary material for "Predicting PANSS symptoms in schizophrenia spectrum disorders using speech only: an international, multi-centre, computational secondary analysis across multiple languages"

**Contents**

Participant recruitment ... 1
Speech elicitation tasks ... 3
Audio preprocessing and feature extraction ... 4
Dataset split ... 4
Machine learning algorithms and pipelines ... 4
Deep learning algorithms and pipelines ... 5
Hyperparameter tuning and model selection ... 6
Learning curves ... 7
Independent dataset for out-of-sample deployment performance ... 7
List of acoustic and prosodic variables ... 7
Parameters of the best models ... 8
References ... 9
Supplementary tables ... 10
Supplementary figures ... 14

### Participant recruitment

Czech-speaking participants were recruited by Filip Spaniel's team from the National Institute of Mental Health (NIMH) of the Czech Republic. The recruitment was approved by the ethics commission of NIMH (No. 179/22). Both inpatients and outpatients were recruited for the study. Inclusion criteria: (1) Males and females aged between 18 and 60 years old; (2) Diagnosed with Schizophrenia (ICD-10 code F20.X), Acute Polymorphic Psychotic Disorder with Symptoms of Schizophrenia (F23·1), or Acute Schizophrenia-Like Psychotic Disorder (F23·2); (3) Participants must either be experiencing their first episode of schizophrenia with a disease duration of less than two years from the onset of initial psychotic symptoms, or have had multiple episodes with a total cumulative disease duration not exceeding 1·5 years. Exclusion criteria: (1) Presence of organic mental disorders, mental retardation with an IQ below 80, severe neurological disease (2) Metal implants in the head or face (3) Presence of a cardiac pacemaker.

English-speaking participants were recruited by Sunny X Tang's team at Northwell Health in New York, the United States, from 4/21/2022 to 10/10/2025, and were part of the PsyCL study.[1] The recruitment was approved by the IRB at Feinstein Institutes for Medical Research, Northwell Health (No. 22-0282-ZHH). The participants were recruited from both inpatient and outpatient units of Zucker Hillside Hospital based on approval from the treatment team. In addition to providing informed consent to participate in the study, participants included in this analysis additionally provided optional consent to share their data with scientists outside of Northwell Health in order to maximize their contribution to science. Inclusion criteriaf: (1) Diagnosis of schizophrenia, schizophreniform disorder, schizoaffective disorder, delusional disorder, unspecified psychotic disorder, bipolar disorder with psychotic features, or major depressive disorder with psychotic features; (2) Fluent in English; (3) Age 18 - 65 years; and (4) Has capacity and willing to sign informed consent. Exclusion criteria: (1) Participants with substance-induced disorders or disorders secondary to a general medical condition were not included as underlying brain changes may differ from "primary" psychiatric disorders; (2) Disorders affecting speech or language, such as aphasia, significant intellectual disability (IQ<70), or language disorder, or movement disorders affecting speech like tardive dyskinesia, or physical impairments causing significant distortions to speech; (3) Serious neurological or endocrine disorder or any medical condition or treatment known to affect the brain and/or language, including but not limited to: neurodegenerative disorders, traumatic brain injury with active symptoms, encephalitis, epilepsy; and (4) Cognitive or language limitations, or any other factor that would preclude subjects providing informed consent.

Spanish-speaking participants were recruited by Rosa Ayesa Arriola's team from Valdecilla Research Institute (IDIVAL) in Santander, Spain, between October 2022 and June 2023, and were part of the PAFIP cohort. The recruitment was approved by the local institutional board (No. 2021.119). Inclusion criteria: (1) meeting Diagnostic and Statistical Manual of Mental Disorders (DSM)-IV criteria for brief psychotic disorder, schizophreniform disorder, schizophrenia or schizoaffective disorder. Exclusion criteria: (1) meeting DSM-IV criteria for drug dependence; (2) meeting DSM-IV criteria for mental retardation; and (3) having a history of neurological disease or head injury.

Chilean Spanish-speaking participants were recruited by Alicia I. Figueroa-Barra's team from the University of Chile in Santiago, Chile, between March 2017 and March 2020. These participants were recruited from the outpatient mental health service where they were under regular clinical management. The recruitment was approved by the review board (Ethics Committee for Clinical Research, CEC SSMS of Santiago, Chile) (No. 163-2019). Inclusion criteria were: (1) meeting Diagnostic and Statistical Manual of Mental Disorders (DSM-IV) criteria for brief psychotic disorder, schizophreniform disorder, schizophrenia, or schizoaffective disorder. Exclusion criteria were: (1) personal history of medical and/or neurological disorders, and (2) current alcohol/substance abuse. All participants included in this study from this dataset were first-episode psychosis patients.

French-speaking participants were recruited by two psychologists (Barthelemy S. and Lecomte A.) from Maxime Amblard and Michel Musiol's team from Université de Lorraine. Inpatients or outpatients or in treatment programs were recruited by the Department of Psychiatry in Montperrin Hospital, CH Aix-en-Provence, France, over a period of five months. The recruitment was approved by the ethical commission of the hospital (No. 2015-A01457-42). The dataset is part of the corpus collected within Mopal-D project (Psychopathological Modeling through the Interactional and Linguistic Approach to the Interpretative Mechanism in Psychotic Patients. Therapeutic Proposals), a joint work program between Montperrin Hospital, Aix-Marseille University, and the CNRS (the French National Center for Scientific Research, Joint Research Unit 7118 Atilf). Inclusion criteria: (1) meeting the criteria of the Diagnostic and Statistical Manual of Mental Disorders (DSM-IV) for schizophrenic or paranoid disorders presenting with delusions; (2) aged over 18. Exclusion criteria: (1) lack of capacity to give consent.

Swiss German-speaking participants were recruited by Philipp Homan's team from the Psychiatric University Clinic Zurich and University of Zurich in Zurich, Switzerland, between April 2021 and March 2024. This dataset was collected within the cross-sectional research project, Ventral language stream in schizophrenia with regard to semantic and visuo-spatial processing anomalies (VELAS), and was previously reported in several papers.[3–5] The recruitment was approved by ethics committee of Kantonale Ethikkommission Zurich (KEK-ZH 2020/01049). Both inpatients and outpatients with early psychosis were recruited. Inclusion criteria: (1) met ICD-10 criteria for psychotic disorders; (2) between 14 and 40 years old and within the first eight years of psychosis onset; (3) right-handed, as assessed by the short form of the Edinburgh Handedness Inventory; (4) admitted to the Psychiatric University Clinic Zurich in Zurich, Switzerland. Exclusion criteria: (1) substance intoxication or withdrawal, including cannabis use, for at least one month; (2) ophthalmological or neurological conditions; (3) head trauma; (4) any history of loss of consciousness; and (5) pregnancy.

Dutch-speaking participants were recruited by Iris Sommer's team from UMCG in the Netherlands. The participants were inpatients and outpatients recruited within HAMLETT (ERB number METc 2017/343, EudraCT registration 2017-002406-12) and RAPSODI (ERB number: 16-103, EudraCT registration: 2015-004483-11) RCT cohorts. This dataset has been previously reported in various studies.[6–8] The recruitment was approved by review boards of the University medical center Utrecht (for RAPSODI trial) and Groningen (for HAMLETT cohort). The participants of RAPSODI RCT were recruited either in the Netherlands (Institute for Mental Health Care Eindhoven (GGzE), Eindhoven; Psychiatric Center Geestelijke Gezondheidszorg Centraal (GGz Centraal), Amersfoort; UMCU, Utrecht; Reinier van Arkel Institute for Mental Health Care (RvA), 's-Hertogenbosch), or in a clinic in Belgium (Ziekenhuis Netwerk Antwerpen, ZNA, Antwerp). In RAPSODI RCT, participants were eligible for inclusion if they were native Dutch speakers of 18 years of age or older and met the Diagnostic and Statistical Manual of Mental Disorders, Fourth Edition (DSM-IV) criteria for psychosis not otherwise specified (NOS), schizophreniform disorder, schizophrenia, or schizoaffective disorder. For inclusion, participants also needed to be able to understand the study and be on a stable dose of antipsychotic medication for at least two weeks. Women of childbearing potential who are sexually active were required to use non-estrogenic contraception during treatment and for four weeks afterward. Female participants with post-coital uterine bleeding had to have a normal Pap smear within the past five years or agree to undergo one. Women aged 52 to 75 needed a normal mammogram within the last two years (as part of the Dutch or Belgian breast cancer screening programme) or be willing to have one if not regularly screened. Among exclusion criteria were uncorrected hearing problems or speech disorders (e.g. stutter). The participants for HAMLETTE study were recruited across 24 sites in the Netherlands, involving academic medical centers and mental health institutions. These included the Universitair Medisch Centrum Groningen (coordinator) and Lentis, Academisch Medisch Centrum Amsterdam, Universitair Medisch Centrum Utrecht, Mondriaan/Universiteit Maastricht, and the TRIMBOS Instituut. Additional collaborating mental health organizations included Arkin, Altrecht, Delta, Dimence, GGNet, Mediant, Pro Persona, Vincent van Gogh, and several regional Geestelijke Gezondheidszorg (GGZ) centers such as Rivierduinen, Ingeest, Eindhoven, NoordHollandNoord, Delfland, Drenthe, Centraal, Breburg, as well as Yulius, Parnassia Groep, and the Reinier van Arkel Stichting. For the HAMLETT study, participants were eligible if they had experienced a first episode of psychosis and were currently using antipsychotic medication, with symptoms in remission for 3–6 months. Additional criteria included age between 16 and 55 years, ability to understand the study and provide written informed consent, sufficient proficiency in Dutch, and participation in no other medication trials besides HAMLETT. Individuals were excluded if they had exhibited dangerous or harmful behavior during the first episode of psychosis (FEP), or if they had received coercive antipsychotic treatment during FEP (based on a judicial ruling).

Turkish-speaking participants were recruited by Emre Bora's team at the Department of Neuroscience and Department of Psychiatry in Dokuz Eylul University in Türkiye. Participants were recruited through the Psychotic Disorders Outpatient Unit of the Department of Psychiatry. All participants were native Turkish speakers. The recruitment was approved by the Ethics Committee of Dokuz Eylul University (No. 8527-GOA). The participants were interviewed using the Structured Clinical Interview for DSM-IV Axis I Disorders and met diagnostic criteria of SSD. Exclusion criteria were personal history of medical or/and neurological disorders and current alcohol/substance abuses. This dataset included 35 first-episode psychosis, 56 chronic schizophrenia patients, and 17 chronic schizoaffective patients.

German-speaking and Chinese-speaking participants were recruited by Alberto Parola's team and collaborators. German-speaking participants were inpatients and outpatients recruited from University of Muenster and the LWL-Hospital Muenster between 2005 and 2016. The recruitment was approved by Ethics Committee of the State Chamber of Physicians Westphalia-Lippe and the University of Muenster (No. 2013-104-f-S & No. 3IIIOhr). The patients had been diagnosed with schizophrenia by experienced psychiatrists using the Structured Clinical Interview for DSM-IV SCID-I. Exclusion criteria: patients with any history of other psychiatric disorders, neurological disorders, serious head injury, alcohol or illegal drug abuse, or insufficient knowledge of the German language were excluded from the study. These samples were reported in several previous studies.[9,10,23-25]

Chinese-speaking participants were recruited from inpatient and outpatient facilities in Renmin Hospital of Wuhan University, from April 2014 to February 2015. The recruitment was approved by the ethics committee of Renmin Hospital of Wuhan University and the Institutional Review Board of the Institute of Psychology, the Chinese Academy of Sciences (No. PH13008). Inclusion criteria: (1) met the diagnostic criteria for schizophrenia according to ICD-10; (2) diagnosed by psychiatrists. Exclusion criteria: (1) a history of severe head trauma or neurological illness or if they had a substance abuse problem according to the ICD-10; (2) estimated premorbid IQ below 70 based on previous history or who were unable to understand spoken Chinese well enough to understand testing instructions. All of the Chinese patients were of Han Chinese ethnicity. These samples were reported in several previous studies.[9–12]

In addition to the table summarizing basic demographics, we additionally provided boxplots and pie plots showing the distribution of demographics here, as shown in Supplementary Figure 1, Supplementary Figure 2, Supplementary Figure 3, and Supplementary Figure 4.

**Speech elicitation tasks**

The Czech, Spanish, Turkish, and part of the Swiss German datasets used the same elicitation protocol including seven tasks (the DISCOURSE protocol, available at www.discourseinpsychosis.org). Melshin et al. desribed this protocol in their study.[13] The protocol included seven tasks in total: (1) free conversation about the participant themselves; (2) free conversation about significant events from recent years (or, if not recalled, events from the past week); (3) free conversation about their health states; (4) describing three Thematic Apperception Test images[14]; (5) describing a multi-panel cartoon with ordered subplots that collectively form a coherent narrative; (6) describing repeated drams (or if no repeated dreams, the most recent dream); and (7) reading the crow story aloud and then immediately recall the story in their own words without looking at the story. Tasks 1–3 were categorized as free speech, tasks 4–5 as picture description, task 6 as dream report, and task 7 was split into reading (reading part) and recall (recall part). This protocol was expected to be finished around 20 – 40 minutes.

Several Swiss German participants described fourteen Thematic Apperception Test images[14] either instead of or in addition to completing the DISCOURSE protocol. These data were categorized into picture description.

The English dataset followed a similar protocol with slightly different prompts. It includes six tasks: (1) free conversation about the participant themselves; (2) describing repeated dreams (or if no repeated dreams, the most recent dream); (3) describing four Thematic Apperception Test images[14], the Cookie Theft picture, and an additional image; (4) describing a multi-panel cartoon with ordered subplots that collectively form a coherent narrative; (5) reading the crow story aloud and then immediately recall the story in their own words without looking at the story; and (6) read a fable out aloud. Tasks 1 was categorized as free speech, task 2 as dream report, tasks 3-4 as picture description, task 6 as reading, and task 5 as split into reading (reading part) and recall (recall part).

The Chilean Spanish, French, and Dutch datasets included clinical interviews, which was categorized as free speech.

In the German and Chinese datasets, participants were asked to watch 12 video clips of around 40 seconds, showing an interaction between animated triangles. After watching each animation, participants were asked to interpret the events. This was categorized as recall.

**Audio preprocessing and feature extraction**

Recordings were manually diarized to remove the interviewer's voice, segmented by task, and further divided into intervals of ≤60 seconds to harmonize length and facilitate downstream applications. Recordings shorter than one minute were left unsegmented. Segments shorter than 10 seconds were excluded from further analysis due to insufficient acoustic content and unstable prosodic features, unless they represented the last segment, in which case they were merged with the preceding one. For example, a 244-second sample yielded three 60 s segments and one 64 s segment. For each speech segment, we first extracted 88 features using OpenSmile[15] from the eGeMAPSv02 feature set,[16] which was developed as a minimalistic acoustic parameter set covering voice quality, pitch profile, as well as spectral and cepstral characteristics. This set includes features related to energy, amplitude, spectral balance, and temporal dynamics. An additional set of 31 prosodic variables was extracted using Prosogram[17], providing complementary information on segment duration, F0 contours, rhythm, and intonation patterns. To further capture higher-order acoustic regularities, we employed a large-scale speech representation model, mHuBERT-147, which encodes each segment as a fixed-length embedding. mHuBERT-147 is a pretrained, massively multilingual HuBERT model trained on 90,000 hours of clean, open-license data.[18] It provides speech representations that are aligned across languages and encode rich acoustic and prosodic information, though the interpretation of these embeddings remains opaque.

**Dataset split**

We stratified 6,664 segments into training, validation, and test sets. Splits were based by participant ID, instead of segments, so no individual appeared in more than one set, preventing train–test leakage across segments from the same participant. The split ratio was 80%, 10%, and 10% of participants, yielding 5,229 segments from 358 participants in the training set, 694 segments from 49 participants in the test set, and 741 segments from 46 participants in the validation set. The proportion of segments in each set closely matched the corresponding participant ratios: 78·47% (training), 10·41% (test), and 11·12% (validation). Data split was stratified by language code (i.e. sites) and disease severity. Disease severity was defined by a binary label as having at least two PANSS subscales larger than 3 (not including 3) or not.[19] Then, we did the data split within each language separately. During data splitting, participants from any class represented by only one member are forced into the training set to avoid stratified-split errors. Within each language, participants were first split 80/20 into training set and a temporal pool using stratification on the severity label when class sizes allow, otherwise falling back to a non-stratified split. The temporary pool was then split 50/50 into validation and test under the same "stratify-if-possible, else fallback" rule (if only one participant remains, they go to validation). **Supplementary Table 1** presented the numbers and ratios of participants and segments for all languages.

**Machine learning algorithms and pipelines**

The input features were forwarded to a pipeline with three steps: (1) standardizing every feature to zero mean and unit variance using *StandardScaler*; (2) fit principal component analysis (PCA) on the standardized data, allowing at most $v - 1$ components, where $v$ is the number of features (note: the number of features is always less than number of subjects); and (3) forwarded to a machine learning regressors.

In the PCA step, the number of components was suggested by parallel analyses, following a four-step approach.[20] To obtain a data-driven baseline ("null") against which to compare the empirical explained variance ratios, we performed a Monte Carlo simulation with 100 iterations.[21] In each iteration, we generated a random matrix with the same shape as the standardized data from $\mathcal{N}(0,1)$, fit PCA with the same maximum number of components $v - 1$, and stored the explained variance ratios. We then averaged these explained variance ratios across iterations to form the null explained variance ratio curve (component-wise mean). Then, the explained variance ratios from the actual data were compared to those from the randomly generated data. Components from the real data would be retained, when and only when they explained more variance (i.e. higher ratio) than components from the random data.

We tested 13 machine learning algorithms using the scikit-learn package (version 1·5·2), and XGBoost regressor using the xgboost package (version 2·1·3). If not specified, 42 is the random seed we used to ensure reproducibility in this study. Parameter tuning was carried out with grid search:

1) Ordinary least squares: no parameter to tune.
2) Ridge regression: sklearn.linear_model.Ridge, linear least squares with L2 regularization that shrinks coefficients toward zero and mitigates multicollinearity. The regularization strength $\alpha$ was tuned using the grid $\{10^{-4}, 10^{-3}, 10^{-2}, 10^{-1}, 1{,}10\}$. Other settings followed scikit-learn defaults.
3) Bayesian ridge regression: sklearn.linear_model.BayesianRidge, Byignwayeson version of ridge regression. We tuned the initial regularization parameters over $\{10^{-4}, 10^{-2}, 1{,}10\}$ for lambda (precision of the weights), and also $\{10^{-4}, 10^{-2}, 1{,}10\}$ for alpha (precision of the noise). Other settings followed scikit-learn defaults.
4) Lasso regression: sklearn.linear_model.Lasso, linear model trained with L1 prior as regularizer that promotes sparsity and enables feature selection. The regularization strength α was tuned over $\{10^{-2}, 10^{-1}, 1{,}10\}$. The maximum number of iterations is 5000. Other settings followed scikit-learn defaults.
5) Elastic net: sklearn.linear_model.ElasticNet, linear regression with combined L1 and L2 priors as the regularizer. The regularization strength α was tuned over $\{10^{-2}, 10^{-1}, 1{,}10\}$. The L1 ratio was tuned over {0·1, 0·3, 0·5, 0·7, 0·9}. The maximum number of iterations is 5000. Other settings followed scikit-learn defaults.
6) Huber regression: sklearn.linear_model.HuberRegressor, L2-regularized linear regression model using Huber loss function (quadratic for small residuals, linear for large). The Huber loss function has the advantage of not being heavily influenced by the outliers while not completely ignoring their effect. We tuned the strength of the squared L2 regularization $\alpha$ over $\{10^{-4}, 10^{-3}, 10^{-2}\}$, and epsilon $\varepsilon$ over {1, 1·35, 2}. The maximum number of iterations is 5000. Other settings followed scikit-learn defaults.
7) Support vector regression (SVR): sklearn.svm.SVR, epsilon-support vector regression. We tuned over two kernels, linear and RBF. The regularization parameters were tuned over these grids: $C \in \{10^{-1}, 1{,}10\}$, $\varepsilon \in \{10^{-2}, 10^{-1}, 1\}$. Other settings followed scikit-learn defaults.
8) Random forest: sklearn.ensemble.RandomForestRegressor. A random forest is a meta estimator that fits a number of decision tree regressors on various sub-samples of the dataset and uses averaging to improve the predictive accuracy and control over-fitting. The regularization parameters were tuned over these grids: n_estimators ∈ {50, 100, 200}, max_depth ∈ {5, 10, 20, 30}, max_features ∈ {0·3, 0·5, 'sqrt', 'log2'}, min_samples_split ∈ {5, 10, 20}, and min_samples_leaf ∈ {1, 4, 8}. Other settings followed scikit-learn defaults.
9) Extra trees: sklearn.ensemble.ExtraTreesRegressor. This estimator is very similar to random forest but fits a number of randomized decision trees (a.k.a. extra-trees) on various sub-samples of the dataset and uses averaging to improve the predictive accuracy and control over-fitting. The regularization parameters were tuned over the same grids as random forest regressor: n_estimators ∈ {50, 100, 200}, max_depth ∈ {5, 10, 20, 30}, max_features ∈ {0·3, 0·5, 'sqrt', 'log2'}, min_samples_split ∈ {5, 10, 20}, and min_samples_leaf ∈ {1, 4, 8}. Other settings followed scikit-learn defaults.
10) Gradient boosting: sklearn.ensemble.GradientBoostingRegressor, additive tree boosting (squared-error loss by default) with weak learners regularized by depth and learning rate. The regularization parameters were tuned over these grids: n_estimators ∈ {50, 100, 200}, max_depth ∈ {5, 10, 20, 30}, max_features ∈ {0·3, 0·5, 'sqrt', 'log2'}, min_samples_split ∈ {5, 10, 20}, min_samples_leaf ∈ {1, 4, 8}, and learning_rate ∈ {0·05, 0·1}. They are the same as random forest and extra trees, but with an additional hyperparameter of learning rate. Other settings followed scikit-learn defaults.
11) AdaBoost: sklearn.ensemble.AdaBoostRegressor, additive boosting with decision tree regressors as weak learners (squared-error loss by default). The regularization parameters were tuned over these grids: n_estimators ∈ {50, 100, 200}, and learning_rate ∈ {0·05, 0·1}. Other settings followed scikit-learn defaults.
12) Extreme Gradient Boosting (XGBoost): xgboost.XGBRegressor, gradient tree boosting with additional regularization terms. The following hyperparameters were tuned: n_estimators ∈ {50, 100, 200}, learning_rate ∈ {0·05, 0·1}, max_depth ∈ {5, 10, 20, 30}, min_child_weight ∈ {1, 5, 10}, and gamma ∈ {0·5, 1, 1·5, 2, 5}. Other settings followed the XGBoost defaults.
13) K-nearest neighbors (KNN): sklearn.neighbors.KNeighborsRegressor, a non-parametric, instance-based method predicting from local neighborhoods. The regularization parameters were tuned over these grids: n_neighbors ∈ {3, 5, 7}, weights ∈ {'uniform', 'distance'}, and Minkowski metric parameter p ∈ {1, 2} (1 for Manhattan, and 2 for Euclidean). Other settings followed scikit-learn defaults.
14) Gaussian progress regressor: sklearn.gaussian_process.GaussianProcessRegressor, nonparametric Bayesian regression with kernel-based similarity functions. The following hyperparameters were tuned: kernel ∈ {Constant*RBF, Constant*RBF+White, DotProduct+White, Matern+White}, and alpha ∈ $\{10^{-10}, 10^{-5}, 10^{-3}, 10^{-2}\}$.

**Deep learning algorithms and pipelines**

Multi-layer perceptron (MLP) is a feedforward neural network consisting of an input layer, one or more hidden layers, and an output layer, which learns complex mappings through nonlinear activation functions. We tested two-layer and three-layer MLPs for the PANSS score prediction.

A two-layer MLP consists of an input layer, one hidden layer, and one output layer, with the input layer counted as the zeroth layer. The hidden layer employs a rectified linear unit (ReLU) activation function and batch normalization for nonlinear transformation and regularization. After activation and normalization, we applied the dropout mechanism, which randomly deactivates a proportion of hidden units during training to prevent co-adaptation of neurons and reduce overfitting. For example, with a dropout rate of 0·3, each hidden unit has a 30% chance of being temporarily set to zero at each training step. The output layer is implemented as a linear unit without activation. A three-layer MLP consists of an input layer, two hidden layers, and one output layer, with the input layer counted as the zeroth layer. Similar to the two-layer MLP, each hidden layer was activated by ReLU and then batch normalized, followed by dropout mechanism. Model training was performed for up to 100 epochs on the training set using the AdamW optimizer with root mean squared error (RMSE) loss, while validation performance was continuously monitored. Early stopping was triggered if no improvement was observed for four consecutive epochs. To reduce overfitting, L2 regularization was used in combination with a OneCycleLR learning rate scheduler. The batch size was fixed at 128.

Search of hyperparameters: initial learning rate$\in \{5 \times 10^{-4}, 3 \times 10^{-4}, 10^{-3}, 3 \times 10^{-3}\}$,decay (L2 regularization) $\in \{0, 10^{-3}, 3 \times 10^{-3}\}$, dropout rate $\in \{0·2, 0·3, 0·4\}$. The hidden width of the hidden layer of the 2-layer MLP ranged from 2 to 50. The search grid for the 3-layer MLP was similar, except for the hidden layers, the first-layer dimension $d_1 \in \{2, \ldots, 50\}$, and the second layer $d_2 \in \{1, \ldots, \lfloor d_1/2 \rfloor\}$.

**Hyperparameter tuning and model selection**

For each model, we trained the model using all combinations of parameters as listed above on the training set. Models were trained on the training set and evaluated on the validation set, with the test data never used during tuning. All models were finally tested the model on the test set. Model performance was mainly evaluated by root mean squared error (RMSE), while R-squared provided additional information on goodness of fit. Root Mean Squared Error (RMSE) is the square root of the average squared difference between observed values $y_i$ and predictions $\hat{y}_i$. Lower RMSE indicates better predictive accuracy, and it has the same units as the target variable. Formula as blow:

$$RMSE = \sqrt{\frac{1}{n}\sum_{i=1}^{n}\left(y_i - \hat{y}_i\right)^2}$$

where $n$ is the number of observations.

R-squared ($R^2$) is the coefficient of determination, which represents how much the proportion of variance in the observed values explained by the independent variables in the model relative to a constant-mean baseline. Best possible score is 1 and it could be negative, which means the model is arbitrarily worse than a constant model with R-squared of 0. Formula as blow:

$$R^2 = 1 - \frac{\sum_{i=1}^{n}\left(y_i - \hat{y}_i\right)^2}{\sum_{i=1}^{n}\left(y_i - \bar{y}\right)^2}$$

where $n$ is the number of observations, and $\bar{y}$ is the mean score. Negative R-squared is flagged as poor fit.

Generalization gap was defined as the relative difference between RMSE scores on the train set and, on the validation, or the test set. Take the validation set as example.

$$Generalization\, gap = \frac{validationRMSE - trainRMSE}{trainRMSE}$$

Previous literature defined the generalization gap typically as the absolute difference between validation (or test) and training error (see, e.g., Ballester et al., 2024).[22] However, because we applied the same selection criteria across all eight PANSS items and model performance could vary substantially between items, we adopted a normalized definition to facilitate comparability. A positive generalization gap indicates that the validation error exceeds the training error, reflecting potential overfitting, whereas a negative gap would suggest that the model performs better on the validation set than on the training set, which is usually implausible and points to noise or instability.

To control for potential overfitting, only configurations with a positive validation generalization gap less than 0·5 were retained for further consideration. Within each model-feature combination, the configuration with the lowest validation RMSE was selected. Second, the best configurations retained from the validation stage were evaluated on the independent test set. For each PANSS item, test RMSE was used as the primary criterion, and the final model was chosen as the configuration with the lowest test RMSE while also satisfying the test generalization gap constraint.

**Learning curves**
For non-neural regressors, we computed sample-size learning curves using 5-fold participant-level cross-validation to avoid subject leakage. Within each fold, RMSE was evaluated at five relative training sizes (10%, 30%, 50%, 70%, 100%) using scikit-learn's *learning_curve* function'. For each size, the mean and standard deviation of RMSE across folds were reported and visualized as RMSE versus the number of training examples. For MLPs, we plotted epoch-wise learning curves from the recorded training history. These curves summarize convergence behavior and potential overfitting (e.g., widening train-validation gaps). Learning curves were visualized as in Supplementary Figure 5 – Supplementary Figure 12.

**Independent dataset for out-of-sample deployment performance**
We utilized a pilot set of an ongoing data collection in the Sant Pau hospital at Barcelona, Spain to test how the model performs on out-of-sample. The planned study duration is 18 months, from March 2024 to November 2025. Five patients with treatment-resistant schizophrenia spectrum disorders were recruited from outpatient facilities of Psychiatry Department in Hospital de la Santa Creu i Sant Pau. The study was approved by the Clinical Research Ethics Committee of the Hospital de la Santa Creu i Sant Pau. Inclusion criteria: (1) Patients between 18 - 65 years old; (2) Diagnosis of Schizophrenia following DSM-5 criteria; (3) Patients have to meet TRS criteria recommended by the Treatment Response and Resistance in Psychosis (TRRIP) Working Group Consensus (Howes et al., 2017); (4) Spanish/Catalan fluent speakers. Exclusion Criteria: (1) Intellectual disability; (2) Meet DSM-5 criteria for other mental health disorders but addictive behaviour disorders; (3) Any contraindication to neuroimaging assessment. Funding: European Research Council (ERC-2023-SyG, 101118756 to WH).

**List of acoustic and prosodic variables**
SpeechRate, NrofSafe, SpeechTime, TotNuclDur, TotInternuclDur, TotPauseDur, PropPause, F0MedianInST, PitchMeanST, PitchStdevST, PitchRange, PitchTopST, PitchBottomST, RawF0_p25, RawF0_p50, RawF0_p75, RawF0_mean, PropLevel, Gliss, Rises, Falls, TrajIntra, TrajInter, TrajPhon, TrajIntraZ, TrajInterZ, TrajPhonZ, NuclDurMean, NuclDurStdev, nPVI_nucldur, NuclDurMedian, F0semitoneFrom27.5Hz_sma3nz_amean, F0semitoneFrom27.5Hz_sma3nz_stddevNorm, F0semitoneFrom27.5Hz_sma3nz_percentile20.0, F0semitoneFrom27.5Hz_sma3nz_percentile50.0, F0semitoneFrom27.5Hz_sma3nz_percentile80.0, F0semitoneFrom27.5Hz_sma3nz_pctlrange0-2, F0semitoneFrom27.5Hz_sma3nz_meanRisingSlope, F0semitoneFrom27.5Hz_sma3nz_stddevRisingSlope, F0semitoneFrom27.5Hz_sma3nz_meanFallingSlope, F0semitoneFrom27.5Hz_sma3nz_stddevFallingSlope, loudness_sma3_amean, loudness_sma3_stddevNorm, loudness_sma3_percentile20.0, loudness_sma3_percentile50.0, loudness_sma3_percentile80.0, loudness_sma3_pctlrange0-2, loudness_sma3_meanRisingSlope, loudness_sma3_stddevRisingSlope, loudness_sma3_meanFallingSlope, loudness_sma3_stddevFallingSlope, spectralFlux_sma3_amean, spectralFlux_sma3_stddevNorm, mfcc1_sma3_amean, mfcc1_sma3_stddevNorm, mfcc2_sma3_amean, mfcc2_sma3_stddevNorm, mfcc3_sma3_amean, mfcc3_sma3_stddevNorm, mfcc4_sma3_amean, mfcc4_sma3_stddevNorm, jitterLocal_sma3nz_amean, jitterLocal_sma3nz_stddevNorm, shimmerLocaldB_sma3nz_amean, shimmerLocaldB_sma3nz_stddevNorm, HNRdBACF_sma3nz_amean, HNRdBACF_sma3nz_stddevNorm, logRelF0-H1-H2_sma3nz_amean, logRelF0-H1-H2_sma3nz_stddevNorm, logRelF0-H1-A3_sma3nz_amean, logRelF0-H1-A3_sma3nz_stddevNorm, F1frequency_sma3nz_amean, F1frequency_sma3nz_stddevNorm, F1bandwidth_sma3nz_amean, F1bandwidth_sma3nz_stddevNorm, F1amplitudeLogRelF0_sma3nz_amean, F1amplitudeLogRelF0_sma3nz_stddevNorm, F2frequency_sma3nz_amean, F2frequency_sma3nz_stddevNorm, F2bandwidth_sma3nz_amean, F2bandwidth_sma3nz_stddevNorm, F2amplitudeLogRelF0_sma3nz_amean, F2amplitudeLogRelF0_sma3nz_stddevNorm, F3frequency_sma3nz_amean, F3frequency_sma3nz_stddevNorm, F3bandwidth_sma3nz_amean, F3bandwidth_sma3nz_stddevNorm, F3amplitudeLogRelF0_sma3nz_amean, F3amplitudeLogRelF0_sma3nz_stddevNorm, alphaRatioV_sma3nz_amean, alphaRatioV_sma3nz_stddevNorm, hammarbergIndexV_sma3nz_amean, hammarbergIndexV_sma3nz_stddevNorm, slopeV0-500_sma3nz_amean, slopeV0-500_sma3nz_stddevNorm, slopeV500-1500_sma3nz_amean, slopeV500-1500_sma3nz_stddevNorm, spectralFluxV_sma3nz_amean, spectralFluxV_sma3nz_stddevNorm, mfcc1V_sma3nz_amean, mfcc1V_sma3nz_stddevNorm, mfcc2V_sma3nz_amean, mfcc2V_sma3nz_stddevNorm, mfcc3V_sma3nz_amean, mfcc3V_sma3nz_stddevNorm, mfcc4V_sma3nz_amean, mfcc4V_sma3nz_stddevNorm, alphaRatioUV_sma3nz_amean, hammarbergIndexUV_sma3nz_amean, slopeUV0-500_sma3nz_amean, slopeUV500-1500_sma3nz_amean, spectralFluxUV_sma3nz_amean, loudnessPeaksPerSec, VoicedSegmentsPerSec, MeanVoicedSegmentLengthSec,

StddevVoicedSegmentLengthSec, MeanUnvoicedSegmentLength, StddevUnvoicedSegmentLength, equivalentSoundLevel_dBp

**Parameters of the best models**
P1: {'dim': 5, 'dropout': 0.4, 'lr': 0.0005, 'l2': 0.001, 'early_epoch': 35}
P2: {'C': 0.1, 'epsilon': 0.2, 'kernel': 'rbf'}
P3: {'dim1': 24, 'dim2': 10, 'dropout': 0.4, 'lr': 0.001, 'l2': 0.001, 'early_epoch': 33}
N1: {'dim1': 18, 'dim2': 1, 'dropout': 0.3, 'lr': 0.001, 'l2': 0.003, 'early_epoch': 43}
N4: {'dim': 40, 'dropout': 0.4, 'lr': 0.003, 'l2': 0.003, 'early_epoch': 22}
N6: {'dim1': 28, 'dim2': 7, 'dropout': 0.4, 'lr': 0.0003, 'l2': 0.0, 'early_epoch': 34}
G5: {'dim': 40, 'dropout': 0.2, 'lr': 0.0005, 'l2': 0.001, 'early_epoch': 16}
G9: {'dim1': 3, 'dim2': 1, 'dropout': 0.3, 'lr': 0.0005, 'l2': 0.001, 'early_epoch': 58}

P1 was modelled using a two-layer MLP with 5 hidden units, a dropout rate of 0.4, a learning rate of 0.0005, L2 regularization of 0.001, and early stopping at epoch 35. P2 was modelled using an SVR with an RBF kernel (C=0.1, epsilon=0.2). P3 was modelled using a three-layer MLP with 24 and 10 hidden units in the first and second hidden layers, respectively, a dropout rate of 0.4, a learning rate of 0.001, L2 regularization of 0.001, and early stopping at epoch 33. N1 was modelled using a two-layer MLP with 18 and 1 hidden units in the first and second hidden layers, respectively, a dropout rate of 0.3, a learning rate of 0.001, L2 regularization of 0.003, and early stopping at epoch 43. N4 was modelled using a two-layer MLP with 40 hidden units, a dropout rate of 0.4, a learning rate of 0.003, L2 regularization of 0.003, and early stopping at epoch 22. N6 was modelled using a three-layer MLP with 28 and 7 hidden units in the first and second hidden layers, respectively, a dropout rate of 0.4, a learning rate of 0.0003, no L2 regularization, and early stopping at epoch 34. G5 was modelled using a two-layer MLP with 40 hidden units, a dropout rate of 0.2, a learning rate of 0.0005, L2 regularization of 0.001, and early stopping at epoch 16. G9 was modelled using a three-layer MLP with 3 and 1 hidden units in the first and second hidden layers, respectively, a dropout rate of 0.3, a learning rate of 0.0005, L2 regularization of 0.001, and early stopping at epoch 58.

### Supplementary tables

**Supplementary Table 1. Number (ratio) of data in the training, test, and validation sets by languages.**

| | | Participants | | | | Segments | | | |
|---|---|---|---|---|---|---|---|---|---|
| **Language codes** | **Languages** | **Training** | **Test** | **Validation** | **Total** | **Training** | **Test** | **Validation** | **Total** |
| cs | Czech | 12 (75·00%) | 2 (12·50%) | 2 (12·50%) | 16 | 41 (77·20%) | 29 (13·36%) | 16 (9·45%) | 307 |
| de | German | 32 (80·00%) | 4 (10·00%) | 4 (10·00%) | 40 | 32 (75·27%) | 38 (11·31%) | 40 (13·43%) | 283 |
| en | English | 33 (78·57%) | 5 (11·90%) | 4 (9·52%) | 42 | 79 (79·57%) | 54 (12·14%) | 42 (8·29%) | 651 |
| es | Spanish | 38 (79·17%) | 5 (10·42%) | 5 (10·42%) | 48 | 110 (77·30%) | 129 (10·45%) | 48 (12·25%) | 1053 |
| escl | Chilean Spanish | 11 (78·57%) | 2 (14·29%) | 1 (7·14%) | 14 | 21 (75·93%) | 5 (19·44%) | 14 (4·63%) | 108 |
| fr | French | 12 (75·00%) | 2 (12·50%) | 2 (12·50%) | 16 | 48 (78·63%) | 121 (6·07%) | 16 (15·30%) | 791 |
| gsw | Swiss German | 28 (80·00%) | 4 (11·43%) | 3 (8·57%) | 35 | 131 (78·34%) | 52 (15·50%) | 35 (6·15%) | 845 |
| nl | Dutch | 70 (79·55%) | 9 (10·23%) | 9 (10·23%) | 88 | 35 (79·60%) | 36 (10·06%) | 88 (10·34%) | 348 |
| tr | Turkish | 86 (79·63%) | 11 (10·19%) | 11 (10·19%) | 108 | 168 (80·09%) | 243 (8·14%) | 108 (11·77%) | 2064 |
| zh | Chinese | 36 (78·26%) | 5 (10·87%) | 5 (10·87%) | 46 | 29 (70·56%) | 34 (13·55%) | 46 (15·89%) | 214 |
| / | Sum | 358 (79·03%) | 49 (10·82%) | 46 (10·15%) | 453 | 694 (78·47%) | 741 (10·41%) | 453 (11·12%) | 6664 |

**Supplementary Table 2. Best model performance (RMSE) at the participant level (aggregation method: mean).**

| Symptom | Model | Feature | Whole test set | Czech | German | English | Spanish | Chilean Spanish | French | Swiss German | Dutch | Turkish | Chinese |
|---|---|---|---|---|---|---|---|---|---|---|---|---|---|
| P1 | 2-layer MLP | m-HuBERT | 1·530 | 0·855 | 1·456 | 1·810 | 0·913 | 0·784 | 3·216 | 1·368 | 1·803 | 0·801 | 1·926 |
| P2 | SVR | m-HuBERT | 1·111 | 0·582 | 0·959 | 1·779 | 0·630 | 1·004 | 2·674 | 1·526 | 0·793 | 0·720 | 0·508 |
| P3 | 3-layer MLP | m-HuBERT | 1·537 | 0·206 | 0·414 | 1·881 | 0·170 | 0·771 | 3·145 | 0·873 | 1·316 | 1·487 | 2·576 |
| N1 | 3-layer MLP | m-HuBERT | 1·027 | 1·034 | 0·721 | 0·806 | 0·419 | 0·653 | 1·423 | 1·153 | 1·051 | / | 2·708 |
| N4 | 2-layer MLP | AcouPros | 1·435 | 1·216 | 0·697 | 2·049 | 0·544 | 1·193 | 3·237 | 1·067 | 0·974 | / | 2·385 |
| N6 | 3-layer MLP | m-HuBERT | 0·842 | 0·607 | 0·985 | 0·207 | 0·985 | 1·145 | 1·183 | 1·120 | 0·543 | / | 1·154 |
| G5 | 2-layer MLP | AcouPros | 0·869 | 0·485 | 0·522 | 0·465 | 0·564 | 1·477 | 2·457 | 0·851 | 0·462 | / | 0·393 |
| G9 | 3-layer MLP | m-HuBERT | 1·285 | 0·549 | 0·633 | 1·162 | 0·430 | 0·091 | 3·050 | 1·199 | 1·225 | / | 2·898 |

**Supplementary Table 3. Best model performance (RMSE) at the participant level (aggregation method: median).**

| Symptom | Model | Feature | Whole test set | Czech | German | English | Spanish | Chilean Spanish | French | Swiss German | Dutch | Turkish | Chinese |
|---|---|---|---|---|---|---|---|---|---|---|---|---|---|
| P1 | 2-layer MLP | m-HuBERT | 1·527 | 1·000 | 1·382 | 1·847 | 0·869 | 0·762 | 3·220 | 1·350 | 1·808 | 0·818 | 1·886 |
| P2 | SVR | m-HuBERT | 1·107 | 0·516 | 0·945 | 1·787 | 0·619 | 1·001 | 2·704 | 1·507 | 0·786 | 0·714 | 0·484 |
| P3 | 3-layer MLP | m-HuBERT | 1·542 | 0·218 | 0·440 | 1·950 | 0·168 | 0·795 | 3·118 | 0·880 | 1·317 | 1·480 | 2·572 |
| N1 | 3-layer MLP | m-HuBERT | 1·030 | 1·031 | 0·772 | 0·799 | 0·467 | 0·628 | 1·485 | 1·142 | 1·070 | / | 2·562 |
| N4 | 2-layer MLP | AcouPros | 1·430 | 1·258 | 0·647 | 2·047 | 0·544 | 1·212 | 3·180 | 1·067 | 0·996 | / | 2·375 |
| N6 | 3-layer MLP | m-HuBERT | 0·855 | 0·546 | 0·973 | 0·193 | 0·995 | 1·175 | 1·235 | 1·132 | 0·548 | / | 1·299 |
| G5 | 2-layer MLP | AcouPros | 0·882 | 0·520 | 0·693 | 0·482 | 0·552 | 1·522 | 2·453 | 0·837 | 0·437 | / | 0·359 |
| G9 | 3-layer MLP | m-HuBERT | 1·282 | 0·521 | 0·511 | 1·180 | 0·419 | 0·097 | 3·046 | 1·236 | 1·226 | / | 2·877 |

**Supplementary Table 4. Best model performance (RMSE) at the segment level.**

| Symptom | Model | Feature | Whole test set | Czech | German | English | Spanish | Chilean Spanish | French | Swiss German | Dutch | Turkish | Chinese |
|---|---|---|---|---|---|---|---|---|---|---|---|---|---|
| P1 | 2-layer MLP | m-HuBERT | 1·544 | 0·989 | 1·734 | 1·899 | 1·036 | 0·896 | 3·139 | 1·402 | 1·834 | 0·866 | 2·074 |
| P2 | SVR | m-HuBERT | 1·318 | 0·630 | 1·006 | 1·839 | 0·657 | 0·818 | 2·467 | 1·778 | 0·807 | 0·731 | 0·530 |
| P3 | 3-layer MLP | m-HuBERT | 1·487 | 0·285 | 0·398 | 1·948 | 0·218 | 0·626 | 2·993 | 0·939 | 1·337 | 1·462 | 2·911 |
| N1 | 3-layer MLP | m-HuBERT | 1·071 | 1·057 | 0·662 | 0·942 | 0·467 | 0·813 | 1·518 | 1·239 | 1·081 | / | 2·719 |
| N4 | 2-layer MLP | AcouPros | 1·521 | 1·307 | 0·882 | 2·096 | 0·600 | 1·078 | 3·098 | 0·959 | 1·012 | / | 2·391 |
| N6 | 3-layer MLP | m-HuBERT | 0·869 | 0·712 | 0·906 | 0·339 | 0·944 | 1·110 | 1·144 | 0·940 | 0·559 | / | 1·238 |
| G5 | 2-layer MLP | AcouPros | 0·951 | 0·550 | 0·709 | 0·488 | 0·572 | 1·647 | 2·267 | 0·676 | 0·479 | / | 0·436 |
| G9 | 3-layer MLP | m-HuBERT | 1·298 | 0·590 | 0·617 | 1·202 | 0·439 | 0·204 | 2·888 | 1·129 | 1·241 | / | 2·898 |

**Supplementary figures**

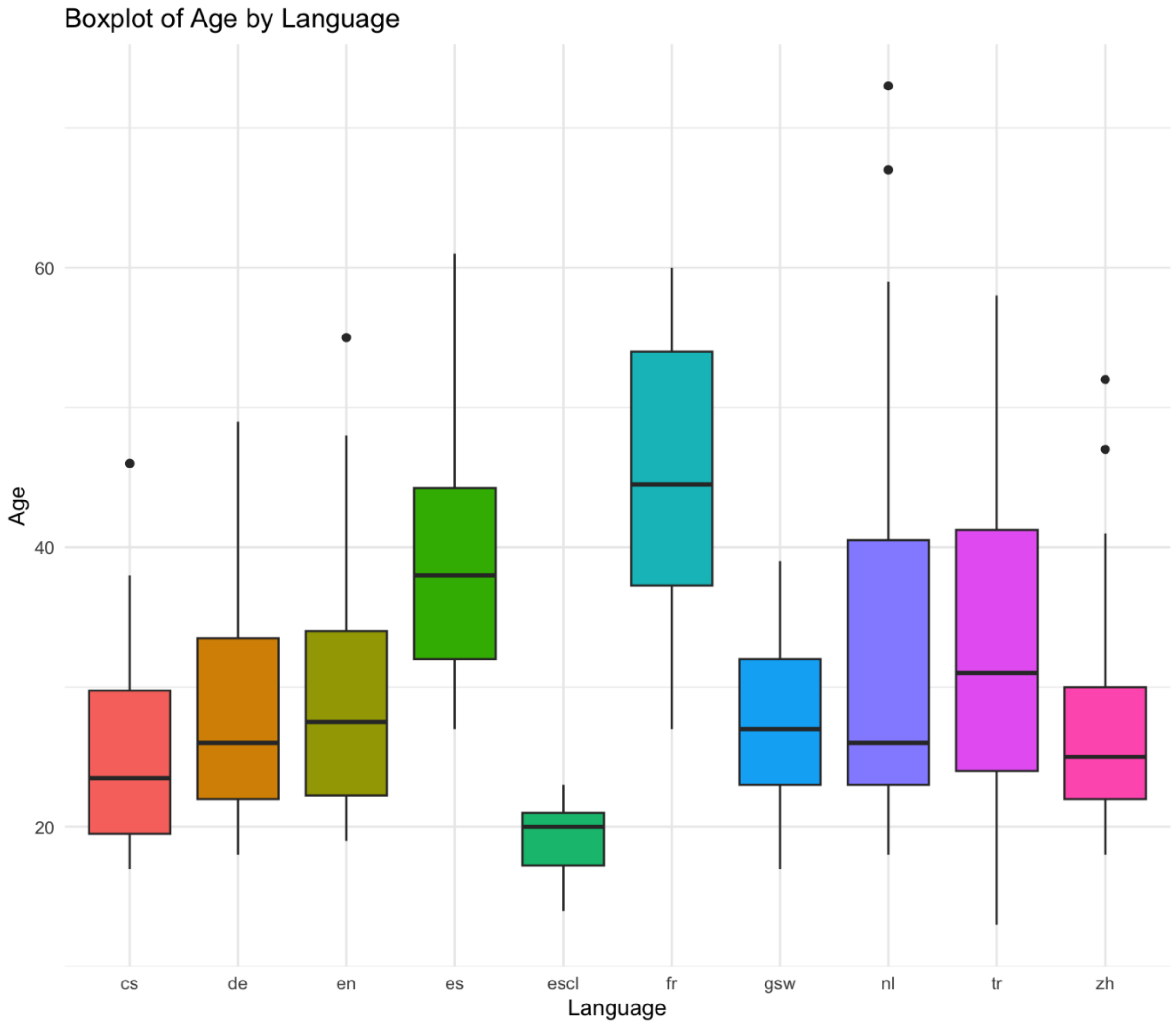


Supplementary Figure 1. Distribution of age in each dataset, as marked by language code: Czech (cs), German (de), English (en), Spanish (es), Chilean Spanish (escl), French (fr), Swiss German (gsw), Dutch (nl), Turkish (tr), and Chinese (zh).

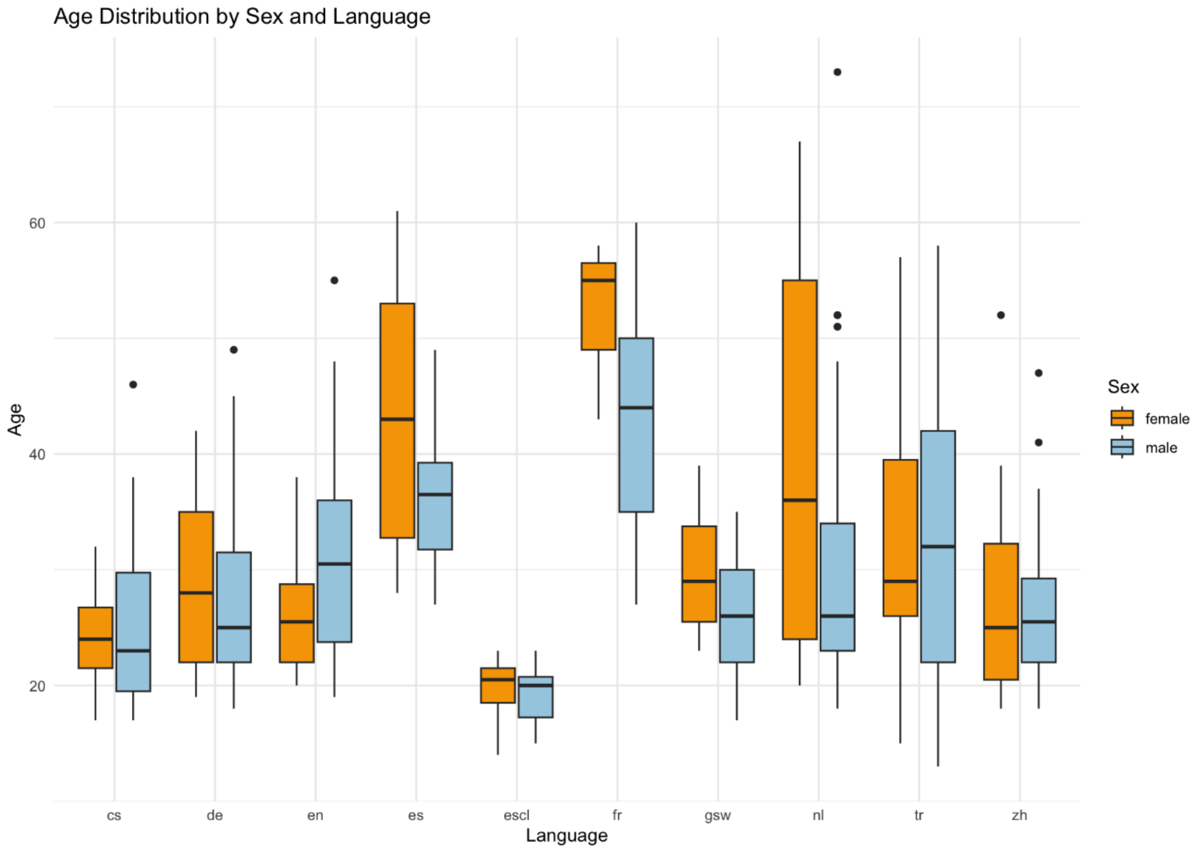


Supplementary Figure 2. Distribution of age by sex in each dataset, as marked by language code: Czech (cs), German (de), English (en), Spanish (es), Chilean Spanish (escl), French (fr), Swiss German (gsw), Dutch (nl), Turkish (tr), and Chinese (zh).

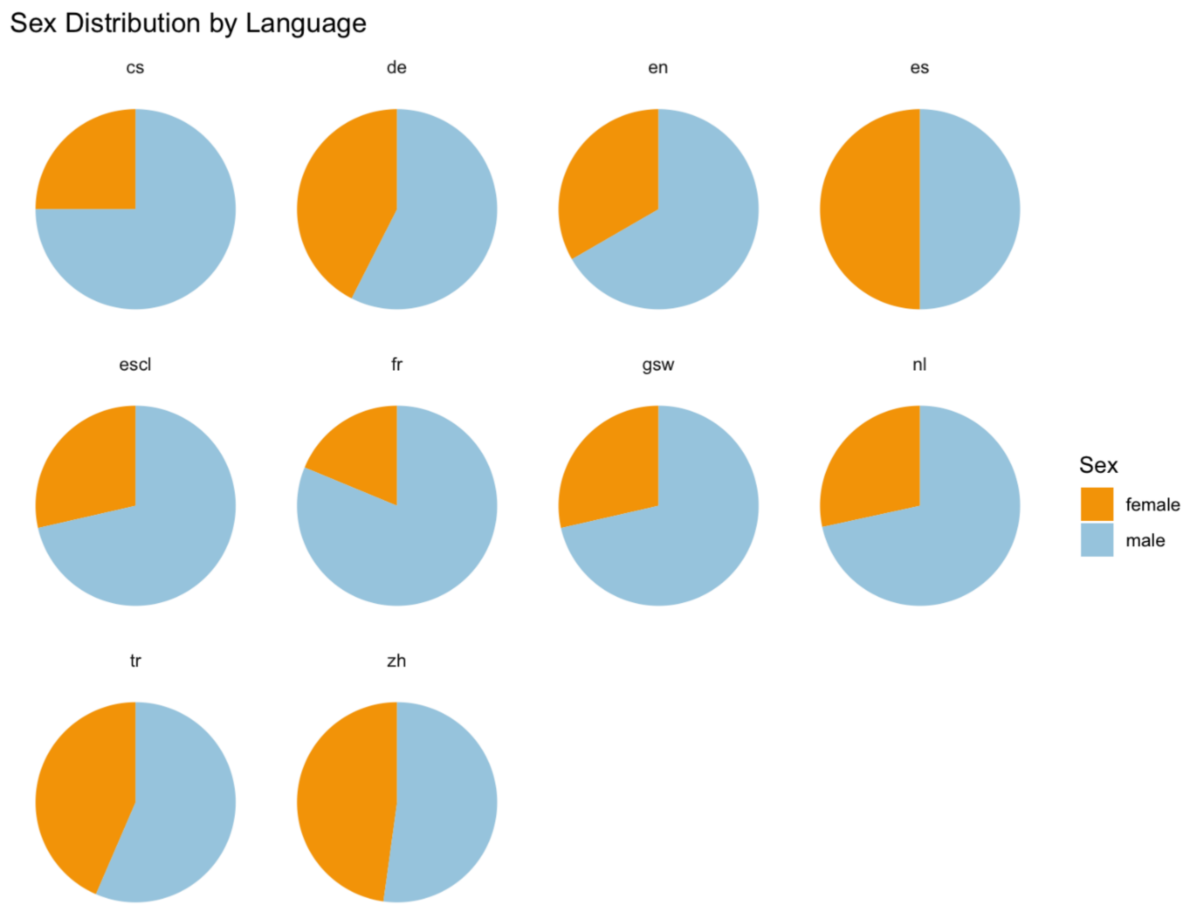


Supplementary Figure 3. Distribution of sex in each dataset, as marked by language code: Czech (cs), German (de), English (en), Spanish (es), Chilean Spanish (escl), French (fr), Swiss German (gsw), Dutch (nl), Turkish (tr), and Chinese (zh).

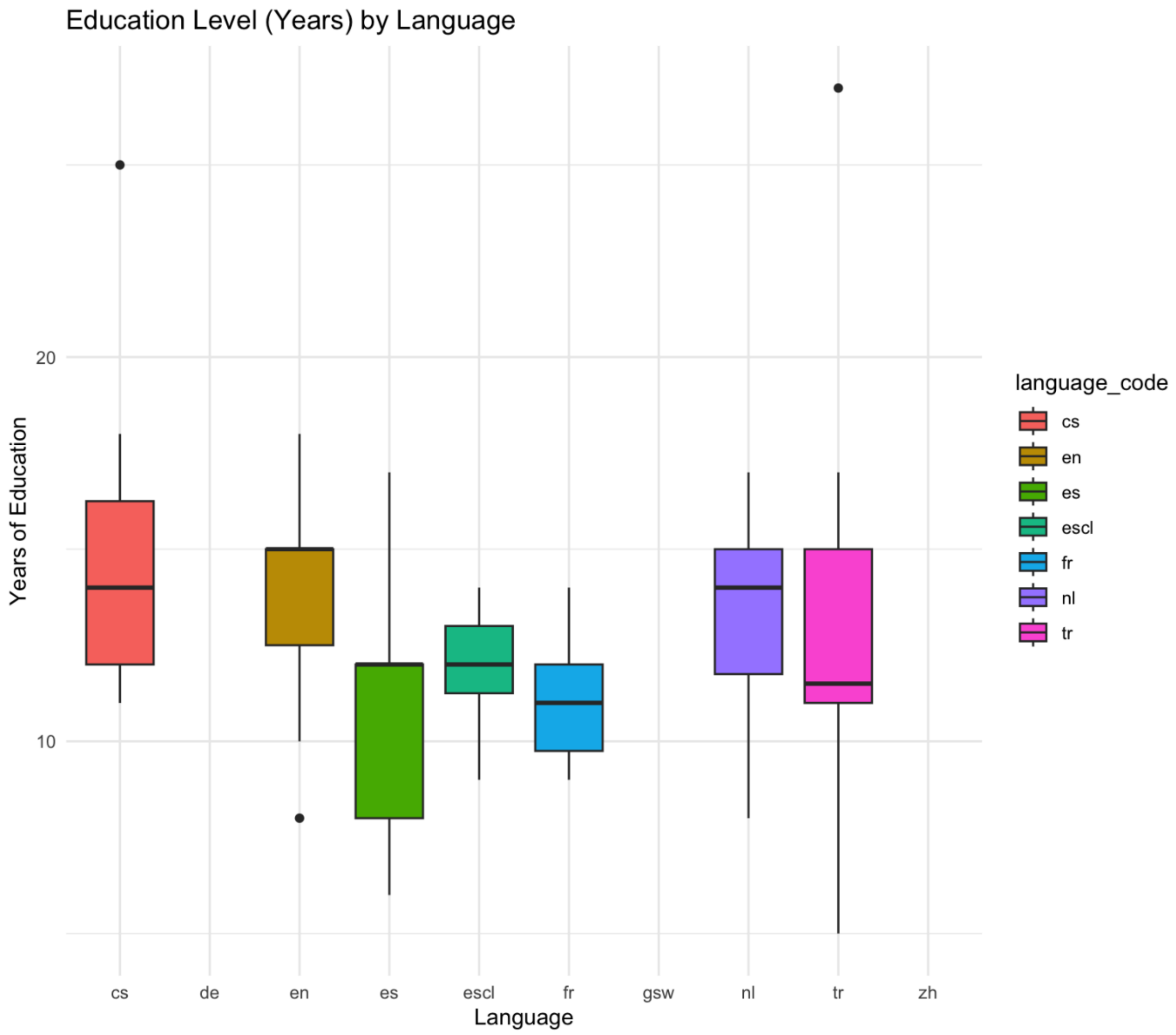

Supplementary Figure 4. Distribution of years of education in each dataset, as marked by language code: Czech (cs), German (de), English (en), Spanish (es), Chilean Spanish (escl), French (fr), Swiss German (gsw), Dutch (nl), Turkish (tr), and Chinese (zh). This data is missing for German (de), Swiss German (gsw), and Turkish (tr) datasets, so there are no boxplots here.

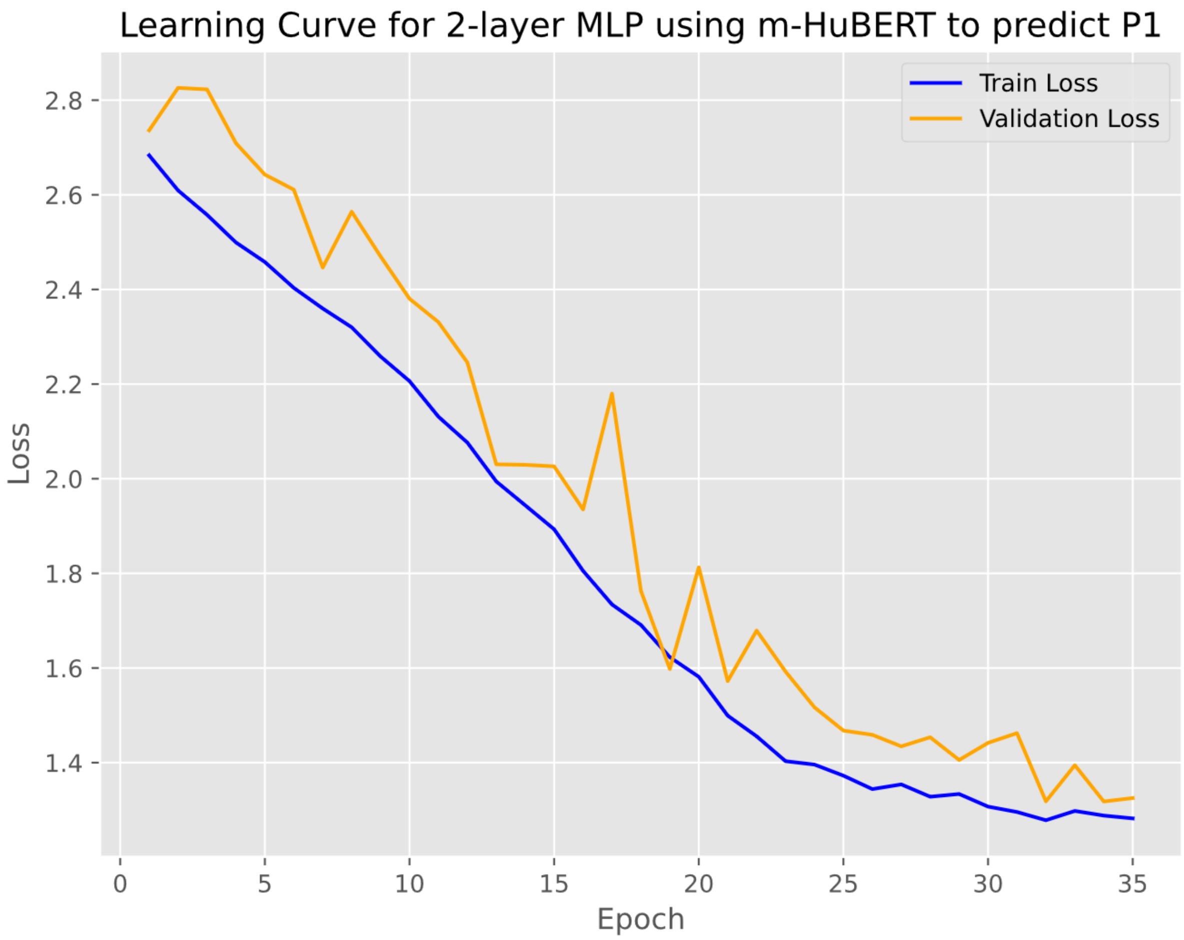


Supplementary Figure 5. Learning curve of P1. Epoch-wise training and validation loss trajectories for the best-performing 2-layer MLP using m-HuBERT features to predict P1. The blue curve shows the training loss and the orange curve shows the validation loss across epochs.

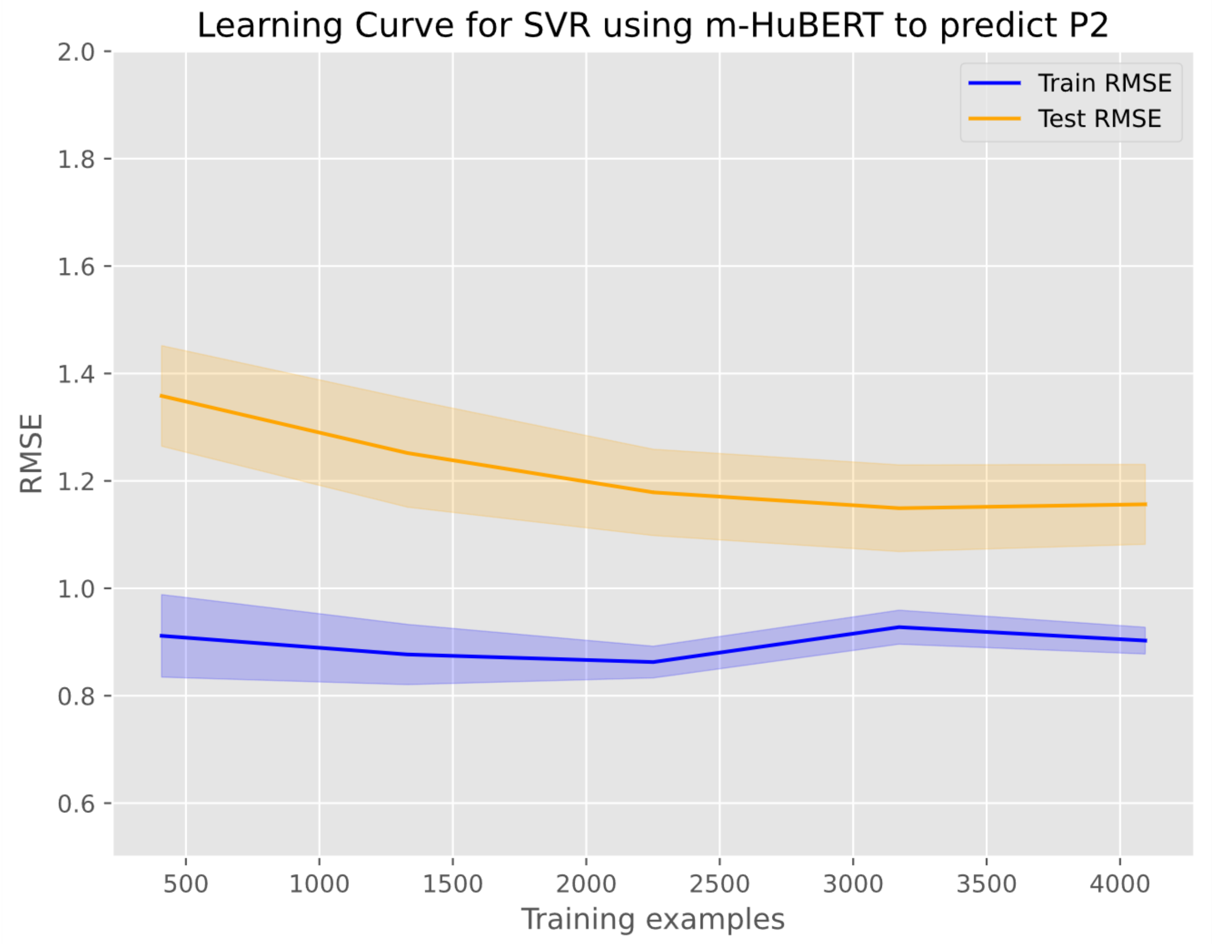


Supplementary Figure 6. Learning curve of P2. Sample-size learning curve for the support vector regressor (SVR) using m-HuBERT features to predict P2. The plot shows root-mean-squared error (RMSE) on the training and validation sets as a function of the number of training examples, with shaded areas denoting standard deviations across 5-fold cross-validation.

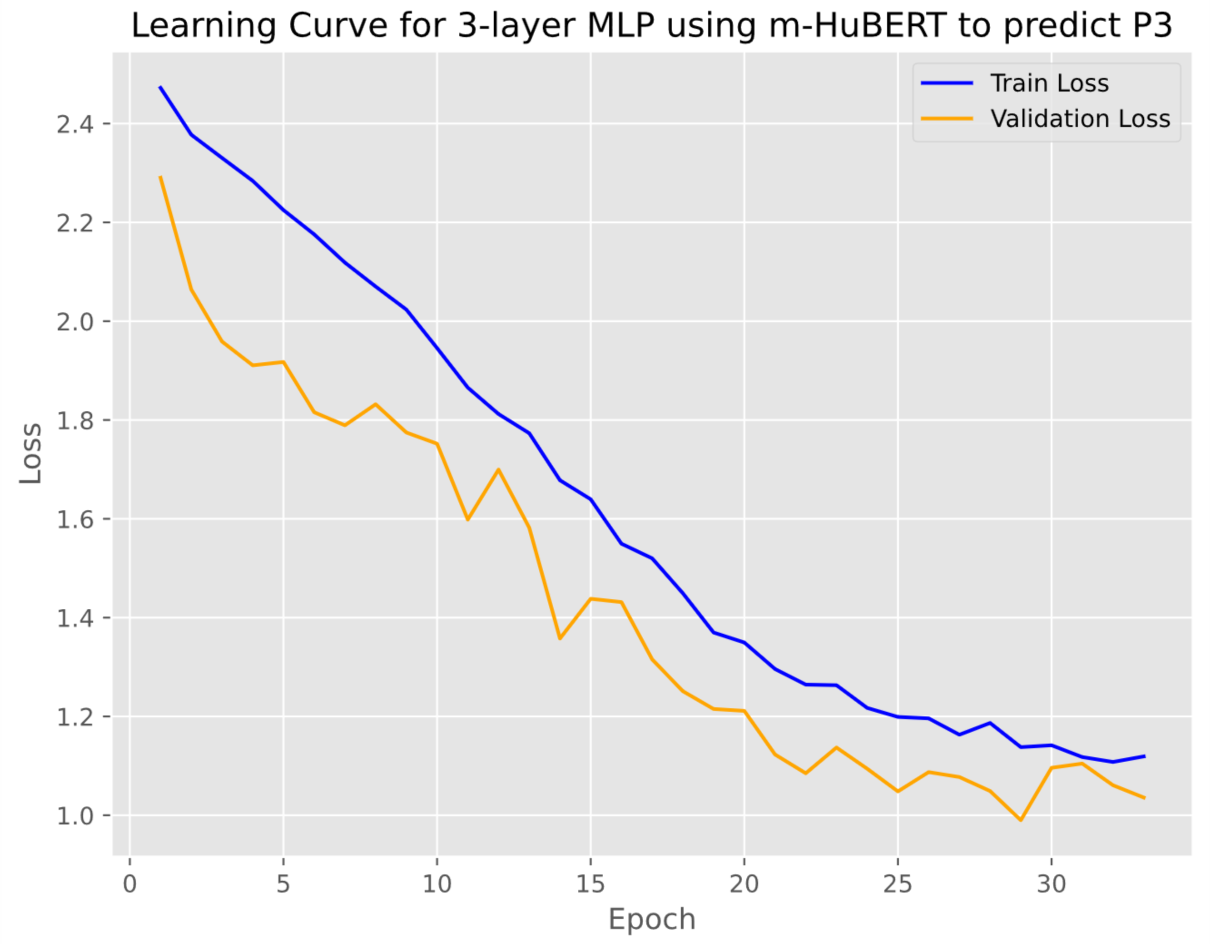


Supplementary Figure 7. Learning curve of P3. Epoch-wise training and validation loss trajectories for the best-performing 3-layer MLP using m-HuBERT features to predict P3. The blue curve shows the training loss and the orange curve shows the validation loss across epochs.

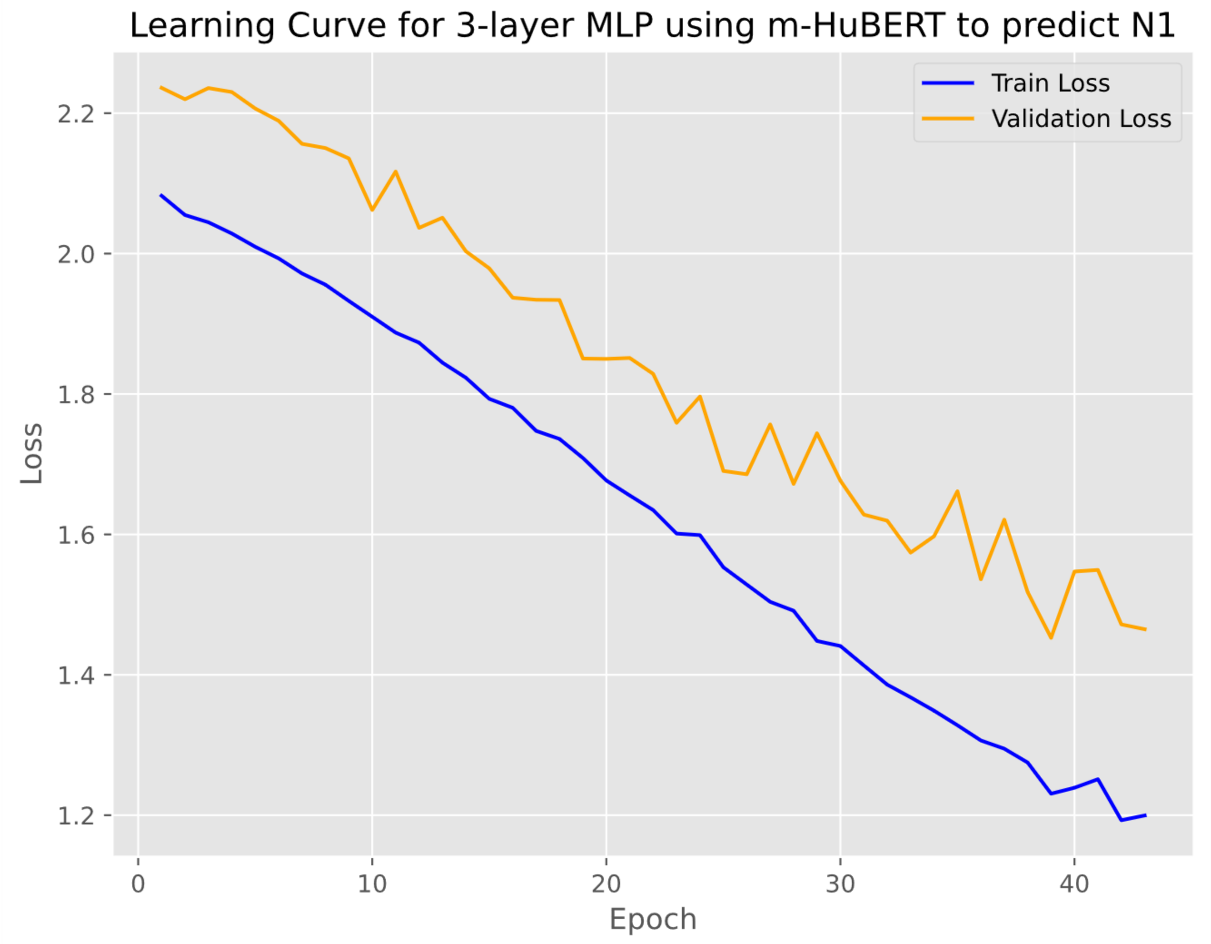


Supplementary Figure 8. Learning curve of N1. Epoch-wise training and validation loss trajectories for the best-performing 3-layer MLP using m-HuBERT features to predict N1. The blue curve shows the training loss and the orange curve shows the validation loss across epochs.

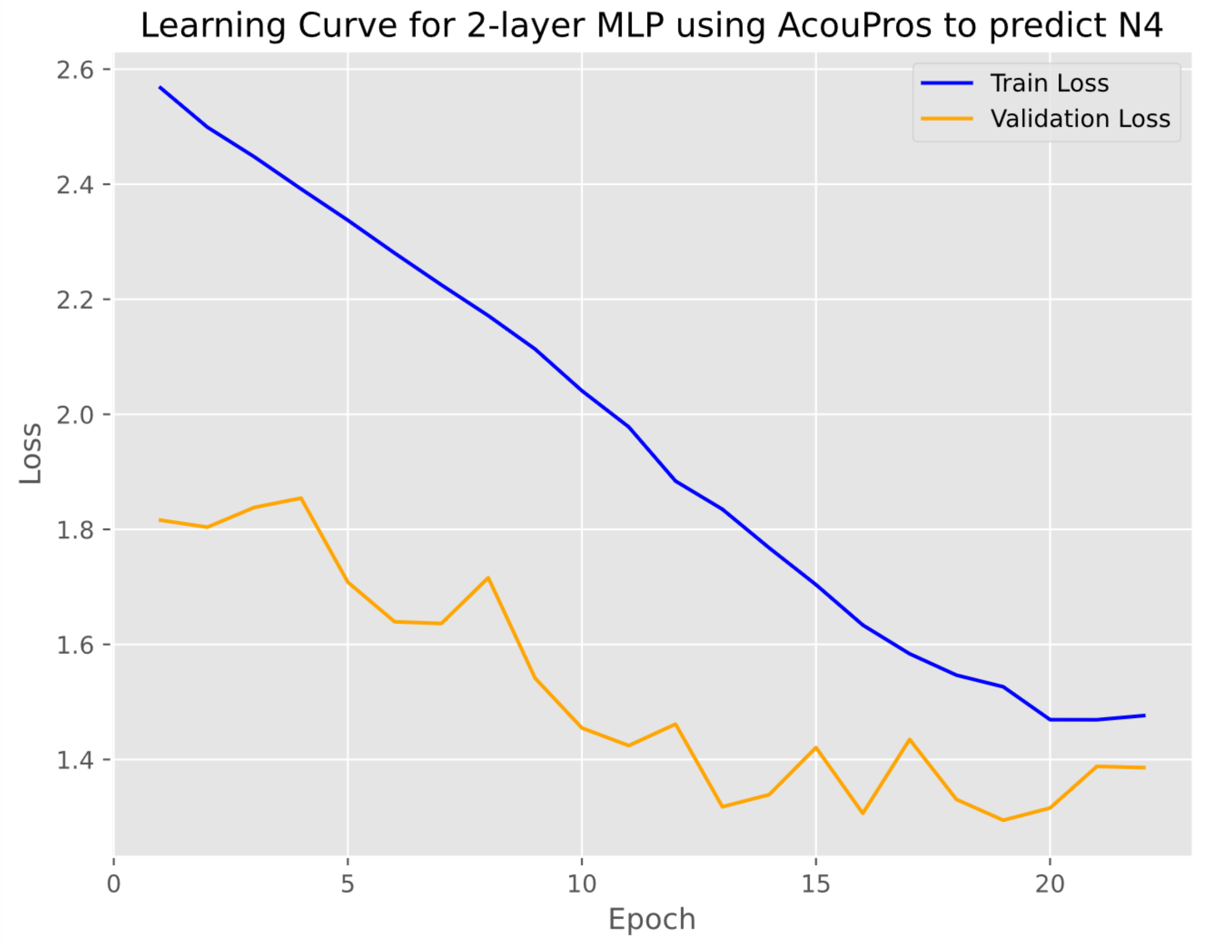


Supplementary Figure 9. Learning curve of N4. Epoch-wise training and validation loss trajectories for the best-performing 2-layer MLP using acoustic-prosodic features to predict N4. The blue curve shows the training loss and the orange curve shows the validation loss across epochs.

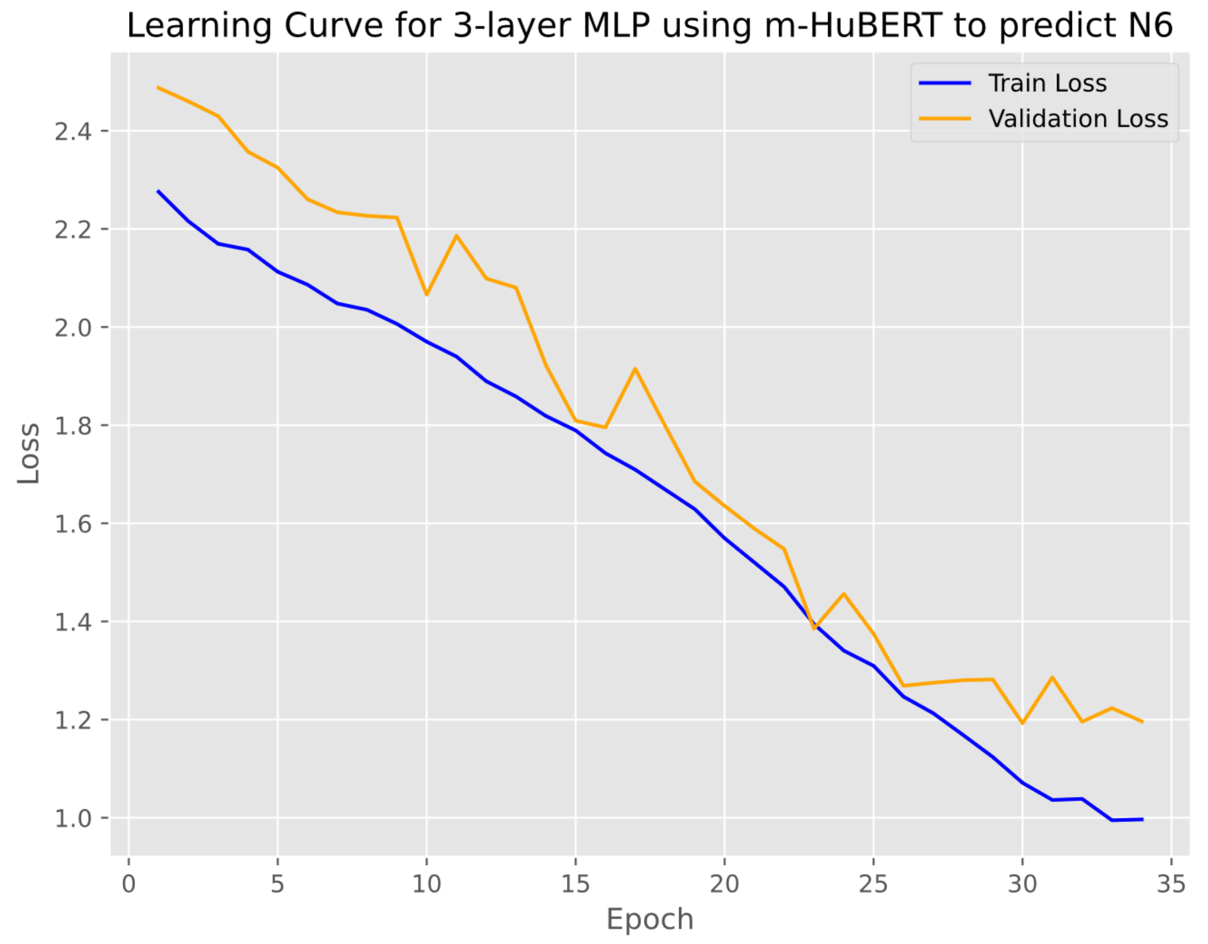


Supplementary Figure 10. Learning curve of N6. Epoch-wise training and validation loss trajectories for the best-performing 3-layer MLP using m-HuBERT features to predict N6. The blue curve shows the training loss and the orange curve shows the validation loss across epochs.

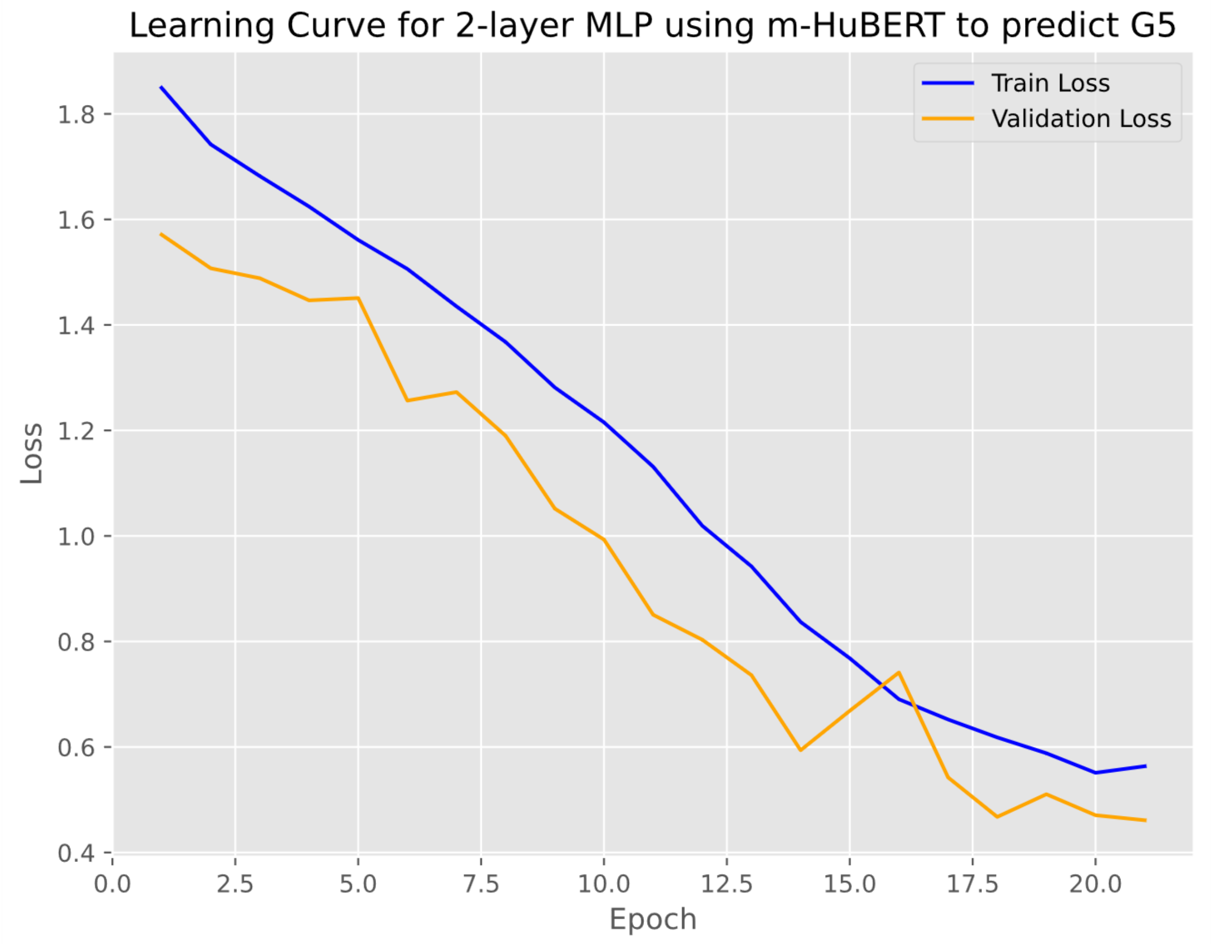


Supplementary Figure 11. Learning curve of G5. Epoch-wise training and validation loss trajectories for the best-performing 2-layer MLP using m-HuBERT features to predict G5. The blue curve shows the training loss and the orange curve shows the validation loss across epochs.

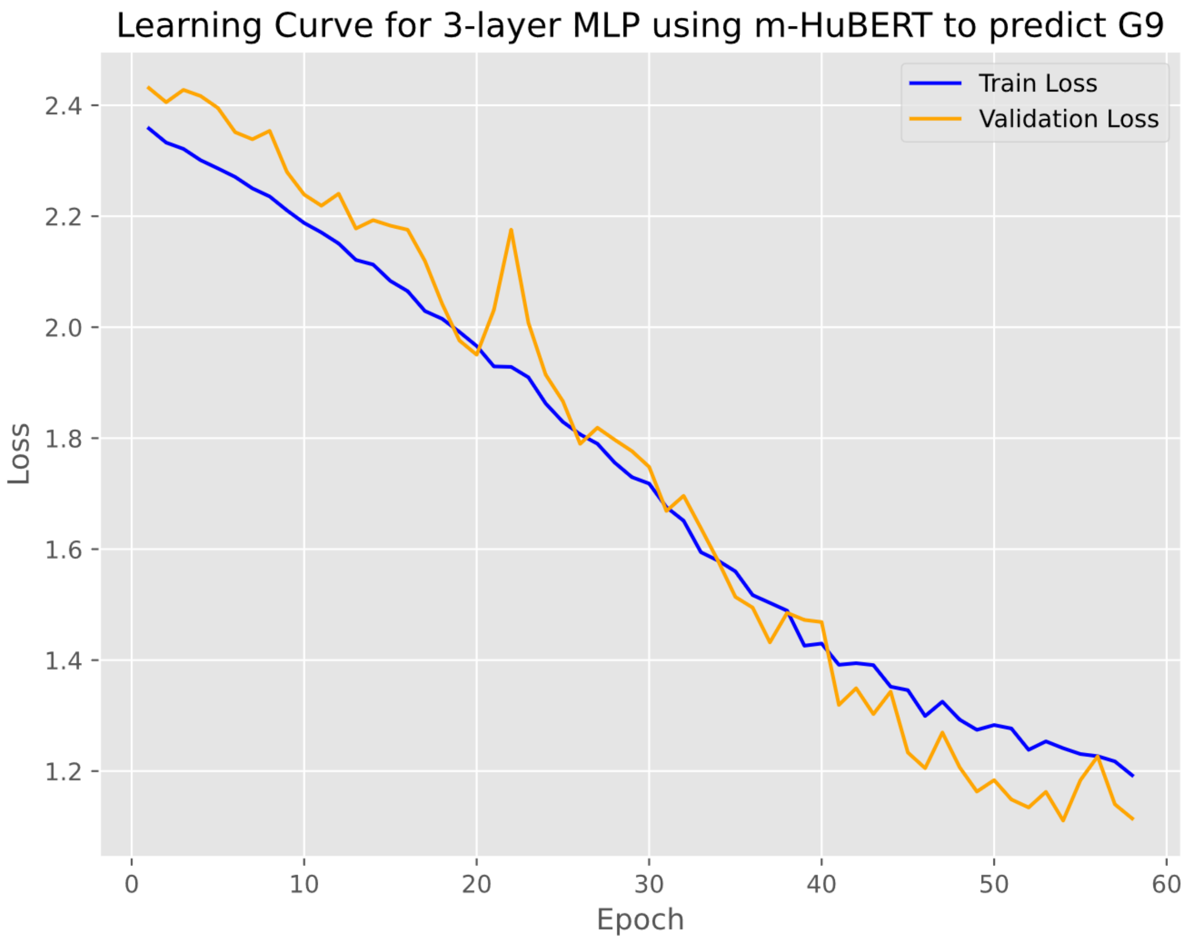


Supplementary Figure 12. Learning curve of G9. Epoch-wise training and validation loss trajectories for the best-performing 3-layer MLP using m-HuBERT features to predict G9. The blue curve shows the training loss and the orange curve shows the validation loss across epochs.

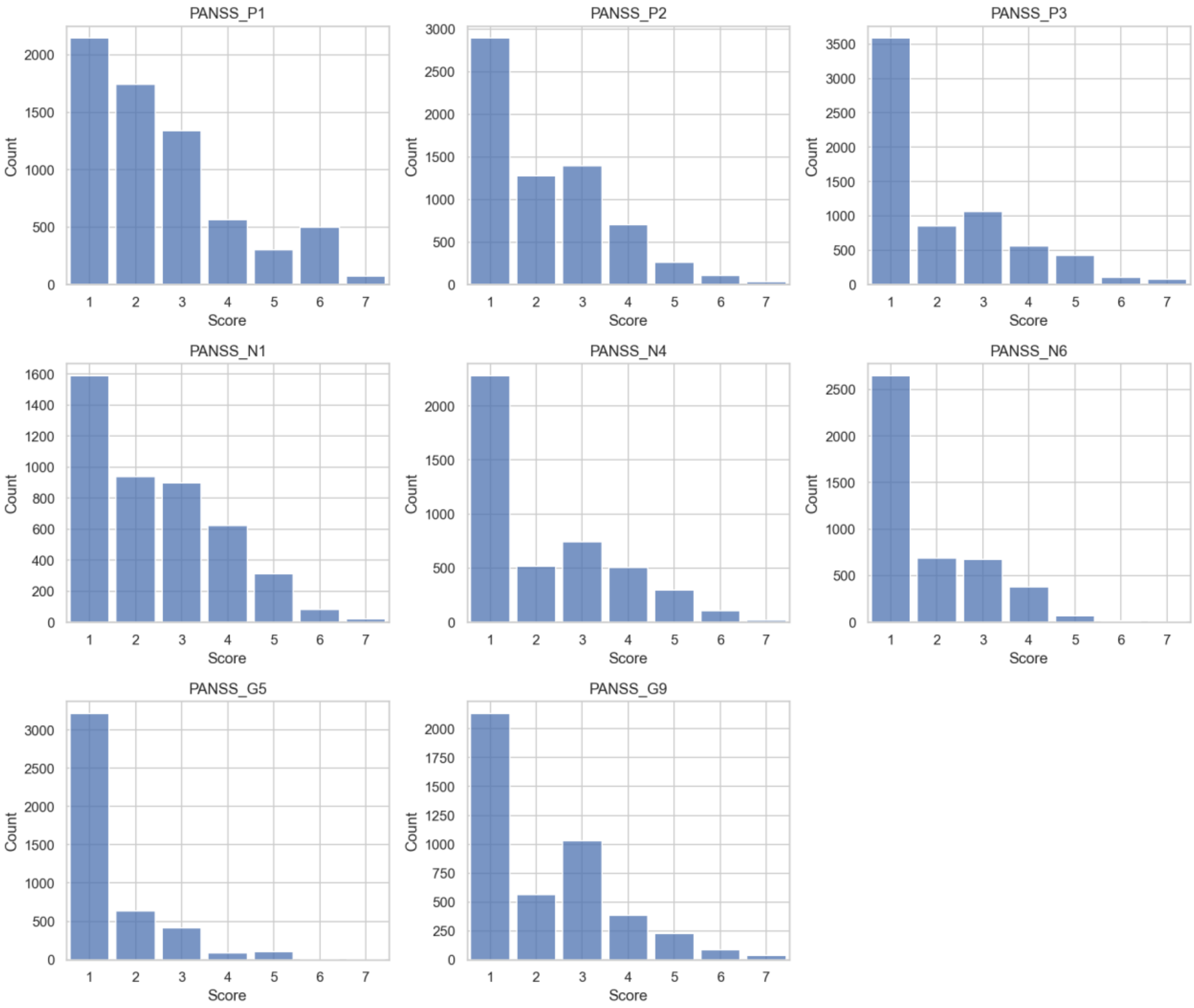

Supplementary Figure 13. Distribution of the PANSS item scores.

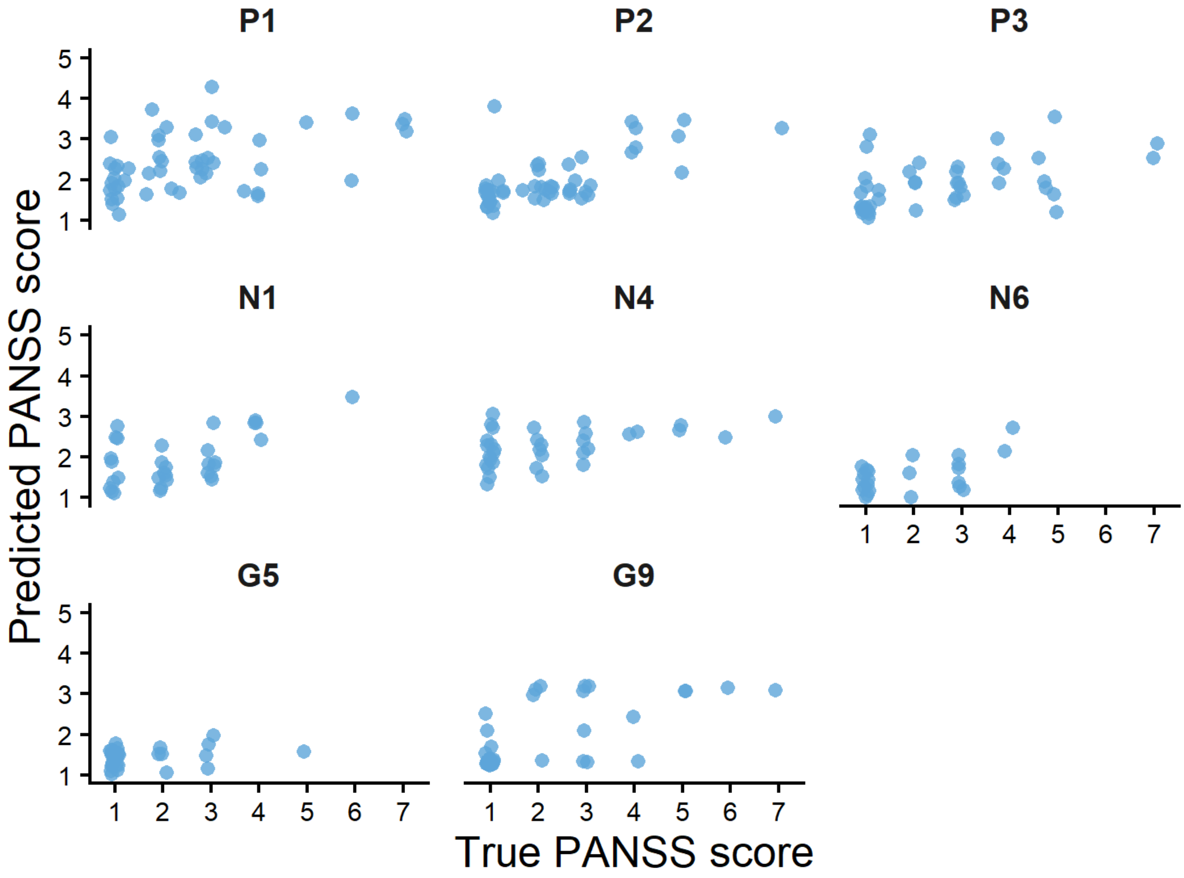

Supplementary Figure 14. Scatter plot of the true and predicted PANSS score.
